# Improving diagnostic performance of kidney allograft rejection with a model combining relative fraction and absolute copies of donor-derived cell-free DNA - results from five independent cohorts

**DOI:** 10.64898/2025.12.01.25341374

**Authors:** Louise Benning, Aylin Akifova, Michael Oellerich, Bilgin Osmanodja, Christian Morath, Julia Beck, Ana Schuetz, Kirsten Bornemann-Kolatzki, Eva V. Schrezenmeier, Thuong Hien Tran, Vedat Schwenger, Maria Shipkova, Eberhard Wieland, Ekkehard Schuetz, Klemens Budde

## Abstract

**Introduction:** Donor-derived cell-free DNA (dd-cfDNA) is a standard-of-care biomarker in kidney transplantation, reported as percentage of total cfDNA or copies/mL. We hypothesized that combining both metrics into a continuous composite score would reduce false classifications thus improving the accuracy of rejection diagnosis.

**Methods:** We analyzed 443 dd-cfDNA measurements in 383 patients from five independent kidney transplant cohorts with both percentage and copies/mL. A random discovery set of 25 biopsy-proven rejections and 26 non-rejections was used to derive a continuous composite score (CM-Score) integrating dd-cfDNA (%) and copies/mL, and validated in 75 rejections and 279 non-rejections for diagnostic performance and compared to 11 published cohorts.

**Results:** For all evaluations, CM-Score was better than cp/mL, which was better than percent. The CM-Score retained a high NPV of 91% (89–93%), while significantly improving PPV to 81% (72–89%; P<0.0001, 25% prevalence), compared to published values (weighted average NPV: 90% (89–91%); PPV: 54% (52–55%), N=6,536). A decision curve analysis yielded a significantly higher net benefit for the CM-Score (P<0.003).

**Conclusions:** The CM-Score is a robust and clinically meaningful tool improving diagnostic accuracy for ruling-out and ruling-in rejection using dd-cfDNA as stand-alone metric with high potential to improve decision-making in kidney transplantation.

## 1 Introduction

Donor-derived cell-free DNA (dd-cfDNA) has become an integral biomarker in kidney transplantation aftercare and standard-of-care in clinical practice, for both for-cause and routine surveillance. Most commercial tests report dd-cfDNA as percentage of total cell-free DNA (cfDNA), which is referred to as relative quantification of dd-cfDNA (dd-cfDNA(%)(1).

Although dd-cfDNA(%) is used to assess the risk of rejection for more than a decade, it has limitations. Because total cfDNA is mainly derived from the recipient, changes in recipient cfDNA directly affect dd-cfDNA(%). Total cfDNA levels can fluctuate for various reasons(2), leading to falsely high or low dd-cfDNA(%) independent of graft health(3). To address this limitation, several studies have evaluated absolute quantification of dd-cfDNA in genomic copies per milliliter of plasma (cp/mL), consistently demonstrating superior performance compared to dd-cfDNA(%) alone(4-6).

However, reporting two distinct values may lead to disagreement, producing divergent results compared to single fixed thresholds. Consequently, parallel testing approaches have been proposed(4, 6, 7), in which an elevated result from either assay would indicate an increased injury risk. The latter was recently also applied in heart transplant recipients(8). Such approaches may improve sensitivity but reduce specificity, as false positives in either percentage or cp/mL would trigger misclassification.

We hypothesized that integrating dd-cfDNA(%) and copies/mL into a continuous combination score would overcome the limitations of single thresholds, improve robustness, and maintain high specificity for rejection diagnosis. This approach may reduce the influence of outliers and better distinguish true rejection from background variation.

## 2 Methods

### 2.1 Study Design

We combined 443 dd-cfDNA results from 383 patients of five independent cohorts(2, 5, 9-12). Details of the patient populations are reported in the original publications. 265 samples were from patients undergoing indication biopsies. BK-virus associated nephropathy (BKVAN samples) included 32 samples with high BK-viraemia defined as >2.5-fold above the American Society of Transplantation (AST) threshold for presumptive BKVAN (>4 log_10_/mL), and 13 cases with biopsy-proven BKVAN. 25 Control samples with unremarkable histology were included. Histological findings (Table 1) were classified per Banff criteria at time of publication. The 146 Stable samples (without biopsy) were from 86 clinically stable kidney recipients, with stable and normal serum creatinine over the last 6 months within a routine surveillance setting. The validation cohort included 88 stable samples, from 44 patients, contributing 2 samples obtained 5 to 16 weeks apart. To assess whether including both samples from these patients introduced bias, we performed 1000 random samplings, selecting one sample per patient. Sensitivity, specificity, positive predictive value (PPV), and negative predictive value (NPV) were calculated for each iteration. Performance metrics showed no variation (coefficient of variation < 0.001%) compared to the full dataset, confirming that inclusion of paired samples does not bias performance estimates.

**Table 1:**
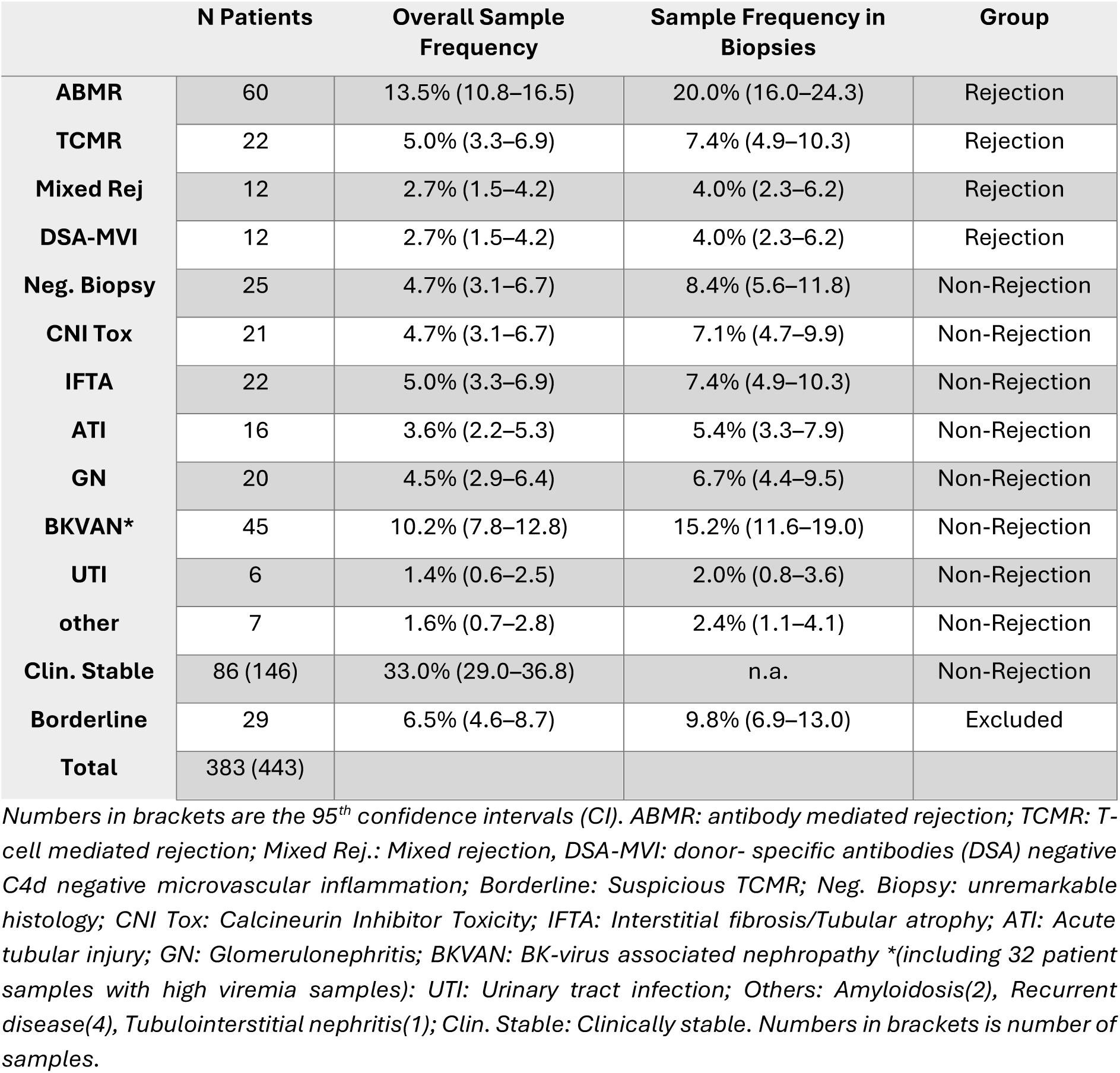
Overview of patient and sample numbers by histopathology.

|  | N Patients | Overall Sample Frequency | Sample Frequency in Biopsies | Group |
| --- | --- | --- | --- | --- |
| **ABMR** | 60 | 13.5% (10.8–16.5) | 20.0% (16.0–24.3) | Rejection |
| **TCMR** | 22 | 5.0% (3.3–6.9) | 7.4% (4.9–10.3) | Rejection |
| **Mixed Rej** | 12 | 2.7% (1.5–4.2) | 4.0% (2.3–6.2) | Rejection |
| **DSA-MVI** | 12 | 2.7% (1.5–4.2) | 4.0% (2.3–6.2) | Rejection |
| **Neg. Biopsy** | 25 | 4.7% (3.1–6.7) | 8.4% (5.6–11.8) | Non-Rejection |
| **CNI Tox** | 21 | 4.7% (3.1–6.7) | 7.1% (4.7–9.9) | Non-Rejection |
| **IFTA** | 22 | 5.0% (3.3–6.9) | 7.4% (4.9–10.3) | Non-Rejection |
| **ATI** | 16 | 3.6% (2.2–5.3) | 5.4% (3.3–7.9) | Non-Rejection |
| **GN** | 20 | 4.5% (2.9–6.4) | 6.7% (4.4–9.5) | Non-Rejection |
| **BKVAN\*** | 45 | 10.2% (7.8–12.8) | 15.2% (11.6–19.0) | Non-Rejection |
| **UTI** | 6 | 1.4% (0.6–2.5) | 2.0% (0.8–3.6) | Non-Rejection |
| **other** | 7 | 1.6% (0.7–2.8) | 2.4% (1.1–4.1) | Non-Rejection |
| **Clin. Stable** | 86 (146) | 33.0% (29.0–36.8) | n.a. | Non-Rejection |
| **Borderline** | 29 | 6.5% (4.6–8.7) | 9.8% (6.9–13.0) | Excluded |
| **Total** | 383 (443) |  |  |  |

Numbers in brackets are the 95<sup>th</sup> confidence intervals (CI). ABMR: antibody mediated rejection; TCMR: T-cell mediated rejection; Mixed Rej.: Mixed rejection, DSA-MVI: donor- specific antibodies (DSA) negative C4d negative microvascular inflammation; Borderline: Suspicious TCMR; Neg. Biopsy: unremarkable histology; CNI Tox: Calcineurin Inhibitor Toxicity; IFTA: Interstitial fibrosis/Tubular atrophy; ATI: Acute tubular injury; GN: Glomerulonephritis; BKVAN: BK-virus associated nephropathy \*(including 32 patient samples with high viremia samples); UTI: Urinary tract infection; Others: Amyloidosis(2), Recurrent disease(4), Tubulointerstitial nephritis(1); Clin. Stable: Clinically stable. Numbers in brackets is number of samples.

Biopsy decisions were made by treating physicians, yielding a real-world cohort spanning early and late post-transplant complications, including not only early delayed graft function (DGF), or borderline/T cell-mediated rejection (TCMR), but also late complications such as antibody-mediated rejection (ABMR) and drug toxicity (e.g., calcineurin-inhibitor (CNI) toxicity); the wide biopsy time range distinguishes this study from other studies focused mainly on the first post-transplant year. (Supplementary Figure S1).

Blood samples were obtained 0-3 days before biopsy at least 2 weeks post engraftment. dd-cfDNA was quantified using two versions(5, 13, 14) of a single-nucleotide polymorphism (SNP)-based assay, leveraging droplet-digital PCR (ddPCR) (Supplementary Table S1). Both methods have been validated to show equivalence (Supplementary Figure S2).

### 2.2 Combination of percent and copies/ml

We assessed correlation (collinearity) between cp/mL and dd-cfDNA percentage in the interval from LLoQ to 2x upper limit of normal (0.5% and 50 cp/mL)(5)). No significant correlation was found (P=0.12, Supplementary Figure S3).

Linear, logarithmic, and quadratic models were evaluated in a discovery cohort of biopsy-proven rejection (N=25) and non-rejection cases (N=26), using AUCROC as the performance endpoint. For randomization, each sample was assigned a random number between 1 and n for the group of rejections and the group of non-rejections.

Samples with numbers ≤25 were used in the derivation cohort. The cohort size met recommended standards for developing a two-variable model, exceeding the minimum of 20 cases per group based on the rule of at least 10 cases per variable(15-17). Excel Solver function was used to find the divisors and exponents for the models with the goal of maximizing AUCROC using rejection vs. non-rejection. The model (Eq.1) achieved the highest observed AUCROC of 0.942 (95% CI 0.890–0.977).

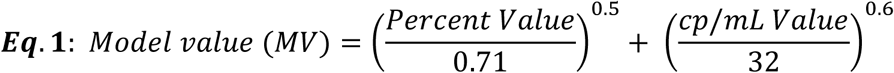

### 2.3 Thresholds

We converted the MV into the “CM-Score” by applying a linear model transformation to set the cut-off to zero (0), resulting in 99% of CM-score values falling into the range of -1 and+2. For percent and cp/mL the prior established cut-offs(5) of 0.5% and 50cp/mL were used for all calculations. The CM-Score was validated in the remaining non-discovery samples, comprising 279 non-rejection and 75 rejection cases as validation group (Supplementary Table S2).

### 2.4 Statistics

Calculations were performed with Python, R, or Microsoft Excel®. Statistical differences are based on non-parametric Mann-Whitney U-test. Confidence intervals (CI) were calculated based on the inverse beta distribution function(18). Likelihood ratios at discrete CM-Score values were defined as the product of the positive (LR^+^) and negative (LR^-^) likelihood ratios at the exact value. For CM-Score, the values between -1 and +1 LR^+^ and LR^-^ were derived from the ROC curve. When the yielded post-test probability values were averaged for values above the cut-off of zero and for values below the cut-off, the correlation of post-test probabilities (prevalences between 5% and 50%) with the 1-negative predictive value (NPV) - below zero and positive predictive value (PPV) - above zero was calculated with R^2^=0.999 and f(x)=1.01x – 0.01, which is within the rounding error of the calculations.

Incremental discrimination was evaluated by calculating bootstrap confidence intervals for the change in AUROC (ΔAUROC) relative to a CM-Score–only reference model.

For the meta-analysis of published data, pooled estimates of sensitivity, specificity, PPV, and NPV were calculated using inverse-variance weighting(19). Pooled estimates were calculated as weighted means with inverse-variance weighting (wi = 1/SEi\), and standard error SE = √(1/Σwi). All between-group comparisons used two-sample Z-tests for independent proportions(20), with Z = (θ^₁ - θ^₂) / √(SE₁² + SE₂²), where θ^₁ and θ^₂ represent group estimates and SE₁ and SE₂ their standard errors(21). Two-tailed p-values were derived from the standard normal distribution. Resulting sensitivity/specificity estimates with CIs were used for decision curve analysis(22, 23).

### 2.5 Methods against bias

Although the model was optimized for AUCROC, results are reported using sensitivity- and specificity-based metrics; the well-understood general difference is shown in the results and discussed for clarity.

The random discovery/validation split was set to 1:6, which avoids overfitting compared to a 1:1 or higher split e.g. 3:1). To address potential overfitting and split bias, we performed complementary validations on the pooled dataset (N=414: 106 rejection, 308 controls): bootstrap validation with 1,000 iterations assessed optimism by resampling with replacement and comparing bootstrap performance to original performance. Random split validation (50 iterations) evaluated stability by randomly partitioning data into 15% discovery/85% validation sets with stratified sampling.

Agreement analysis compared external validation group results (N=354) with bootstrap estimates, adjusting PPV/NPV to 25% prevalence using Bayes’ theorem. Results show high robustness, allowing generalization (Supplementary Figure S4, Table S3).

Differences in handling borderline TCMR in the literature were addressed as detailed in the supplement; briefly, validation data were pooled with and without borderline TCMR and weighted to the literature ratio (69.9%) to ensure unbiased comparability.

## 3 Results

Univariate data evaluation by variable and clinical groups is shown in Figure 1. The statistical significances are given in Table 2, supporting that dd-cfDNA is strongly associated with rejection as published before (9, 10, 24).

**Figure 1:**
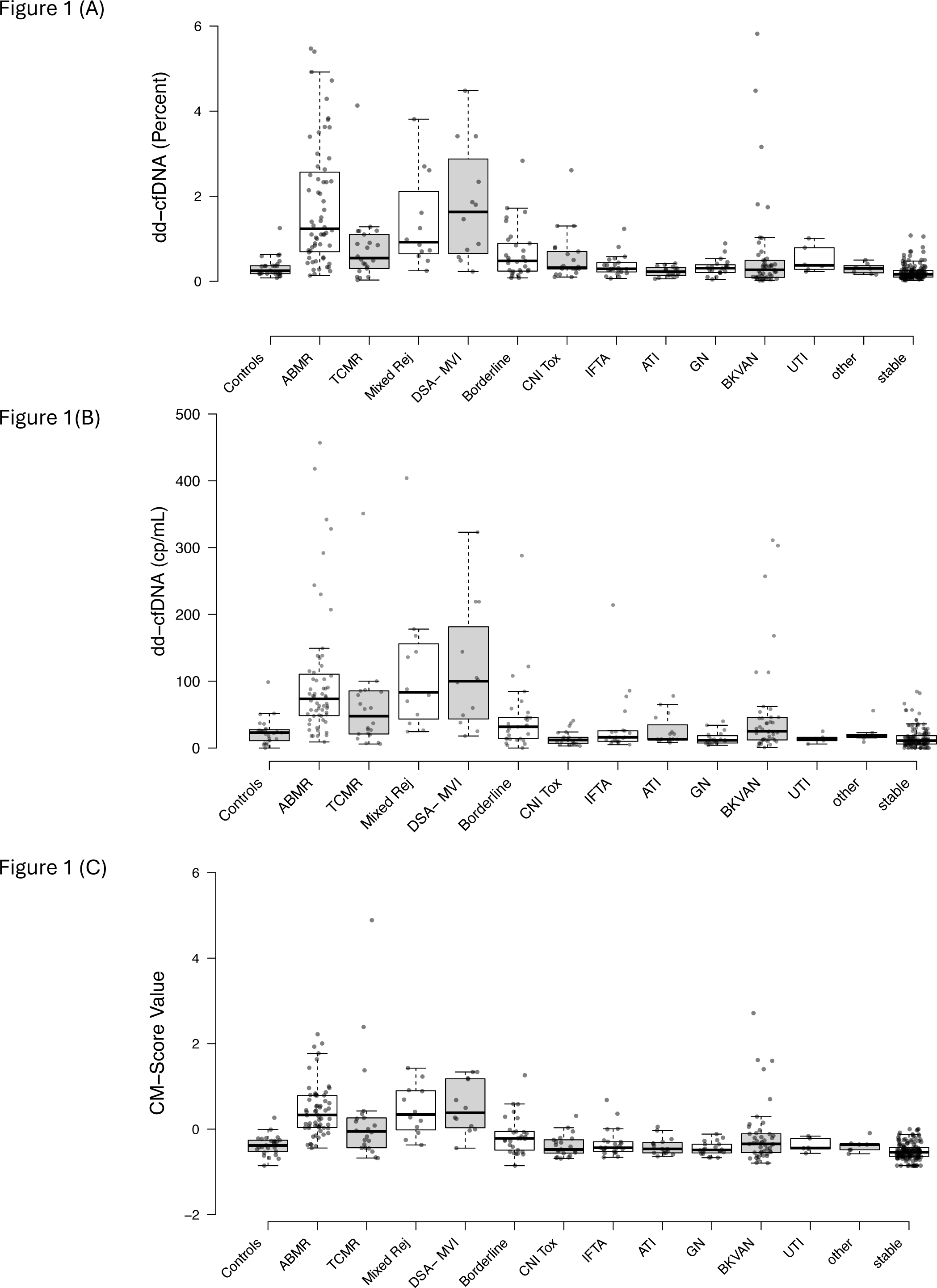
Boxplots of dd-cfDNA in percent (A), cp/mL (B), and the CM-Score (C) segregated by biopsy group. Center lines show the medians; box limits indicate the 25th and 75th percentiles as determined by R software; whiskers extend 1.5 times the interquartile range from the 25th and 75th percentiles; data points are plotted as circles.

**Table 2:** Summary of statistical significances.

| Group | Median % | Median cp/mL | Median CM | U-Test vs. Controls |  |  | U-Test vs. TCMR |  |  |
| --- | --- | --- | --- | --- | --- | --- | --- | --- | --- |
|  |  |  |  | % | cp/mL | CM-Score |  |  |  |
| Controls | 0.25 | 23.0 | -0.40 |  |  |  |  |  |  |
| ABMR | 1.24 | 73.5 | 0.30 | $6.7 \times 10^{-9}$ | $1.82 \times 10^{-8}$ | $1.4 \times 10^{-10}$ | | | |
| TCMR | 0.55 | 47.7 | -0.08 | $4.7 \times 10^{-3}$ | $4.47 \times 10^{-2}$ | $1.4 \times 10^{-3}$ | | | |
| Mixed Rej | 0.92 | 83.5 | 0.31 | $2.5 \times 10^{-5}$ | $3.29 \times 10^{-5}$ | $1.6 \times 10^{-5}$ | | | |
| DSA- MVI | 1.63 | 100.0 | 0.35 | $2.2 \times 10^{-5}$ | $9.53 \times 10^{-5}$ | $1.4 \times 10^{-2}$ | | | |
| Borderline | 0.48 | 31.8 | -0.23 | 0.013 | 0.069 | 0.014 |  |  |  |
| CNI Tox | 0.32 | 12.0 | -0.49 | 0.14 | 0.050 | 0.310 | 0.178 | 0.000 | 0.000 |
| IFTA | 0.29 | 16.0 | -0.45 | 0.27 | 0.407 | 0.391 | 0.021 | 0.006 | 0.006 |
| ATI | 0.23 | 13.0 | -0.48 | 0.14 | 0.426 | 0.235 | 0.002 | 0.008 | 0.008 |
| GN | 0.31 | 11.5 | -0.50 | 0.37 | 0.045 | 0.085 | 0.012 | 0.000 | 0.000 |
| BKVAN | 0.27 | 25.0 | -0.36 | 0.41 | 0.104 | 0.244 | 0.014 | 0.046 | 0.046 |
| UTI | 0.38 | 13.5 | -0.45 | 0.05 | 0.136 | 0.460 | 0.252 | 0.007 | 0.007 |
| other | 0.30 | 18.0 | -0.38 | 0.49 | 0.419 | 0.384 | 0.046 | 0.021 | 0.021 |

CM=CM-Score. Statistical significances of cp/mL, percentage and CM-Score against biopsy proven normal (Controls) and against TCMR. Values for UTI and other are only shown for reference (low numbers).

Across biopsy groups, only the CM-Score distinguished all rejection types from controls and all non-rejection pathologies from TCMR, which is difficult to detect with dd-cfDNA alone (Table 2). In addition, dd-cfDNA increased significantly with TCMR severity (Figure S5).

Figure 2 shows the ROC curves for the validation group. Although the overall AUCROC was not significantly different between the CM-Score and the individual dd-cfDNA metrics, visual inspection of the ROC curve shape suggests that the combination model provides improved diagnostic sensitivity and specificity.

**Figure 2:**
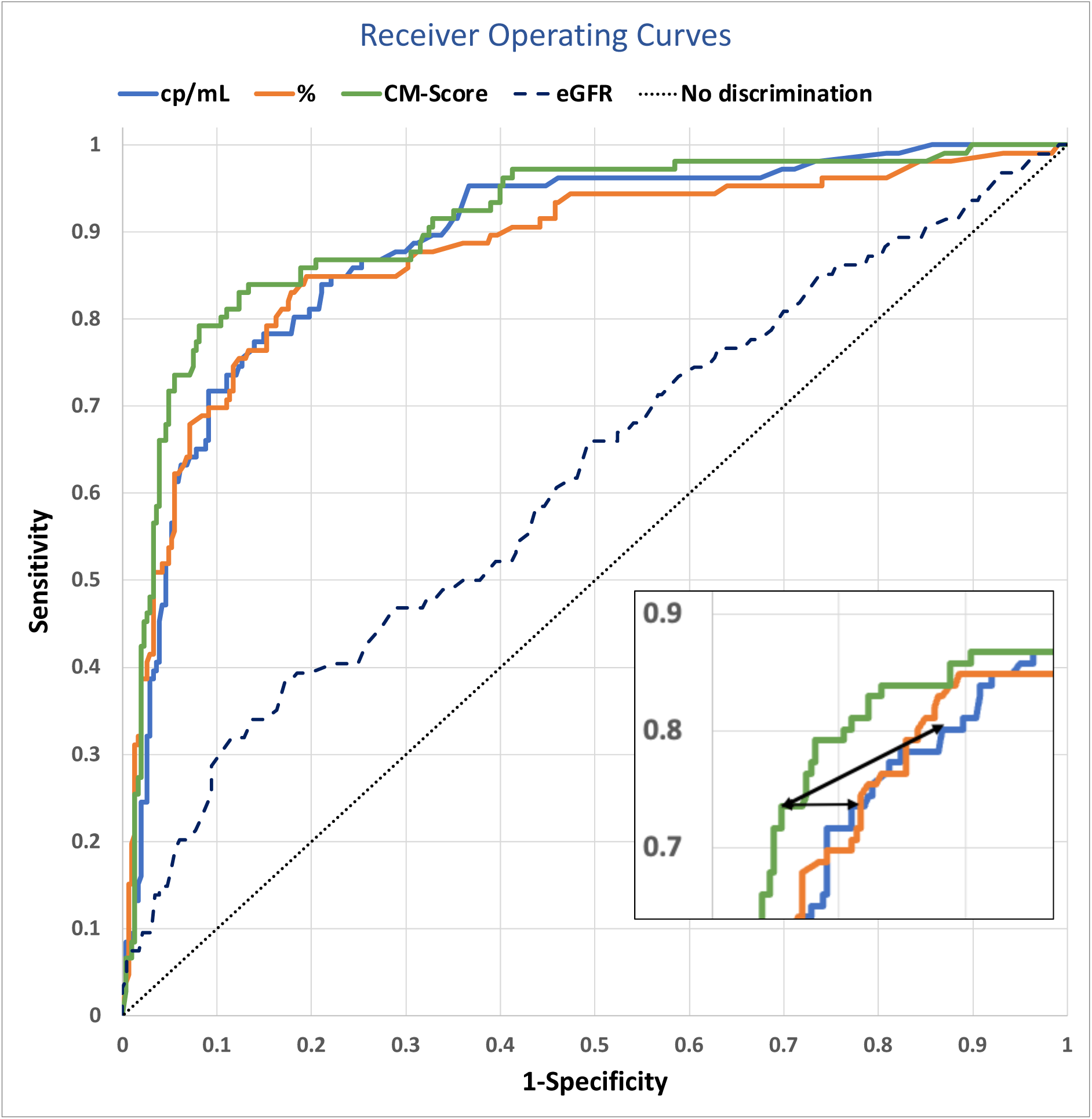
Comparison of the ROC curves obtained from the Validation group (All Biopsy-proven Rejections vs. All Non-Rejections plus Clinically Stable Patients) of the CM-Score, dd-cfDNA percentage, and dd-cfDNA in cp/mL. The insert shows the gain by the CM-Score at the important threshold (depicted by the arrow vs % and cp/mL). The ROC curve for eGFR is shown for reference AUC: 0.62 (0.49–0.73).

dd-cfDNA demonstrated markedly superior performance compared with estimated glomerular filtration rate (eGFR), which showed a sensitivity of 39% (31–48%), a specificity of 82% (77–86%), an AUCROC of 0.62 (0.49–0.73), an NPV of 80% (77–83%), and a PPV of 42% (31–53%). As supplementary metrics, the areas under the precision–recall curves (PRAUC) were 0.76 (0.73–0.79) for the CM-Score, 0.73 (0.71–0.77) for dd-cfDNA percentage, and 0.71 (0.69–0.74) for copies/mL.

### 3.2 Performance Characteristics in the Validation Cohort

We evaluated the most clinically relevant estimators, NPV and PPV, calculated at a prevalence of 25% for the validation cohort. Figure 3 shows the NPV and PPV for CM-Score, percentage, and cp/mL dd-cfDNA. While the NPV remained unchanged, the PPV improved significantly with the CM-Score (P< 0.001). dd-cfDNA(cp/mL) showed a better PPV than dd-cfDNA(%), but the CM-Score outperformed either single metric alone. The full set of performance metrics, including AUC, sensitivity, specificity, PPV, and NPV, is provided in Table 3.

**Figure 3:**
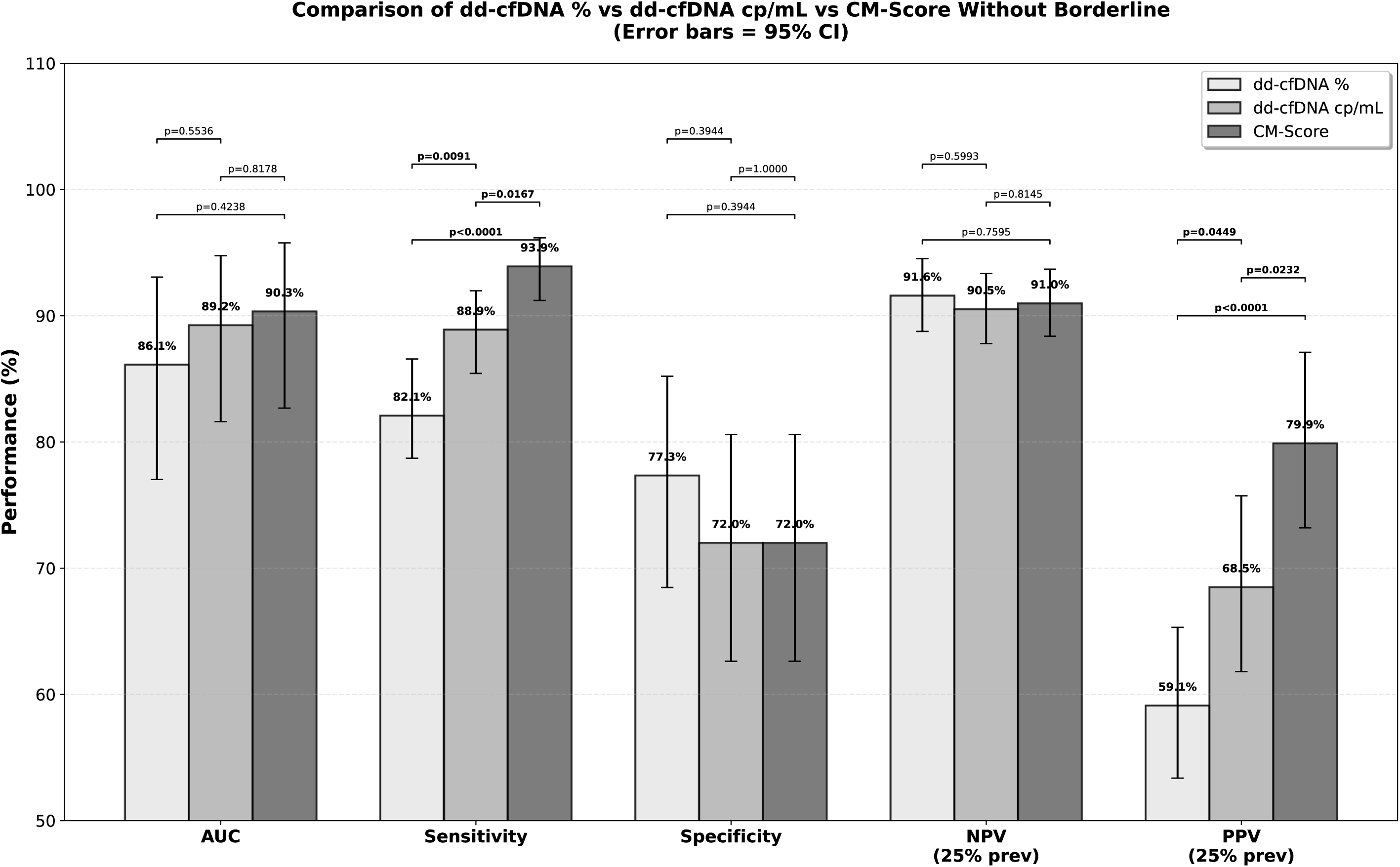
Comparison of performance characteristics. The PPV at 25% prevalence shows a significant improvement to both single values (*P*<0.001) with retained high NPV (not significantly different). This is due to the significant improvement of the specificity with the CM-Score.

**Table 3:** Summary of the performance parameters of the CM-score, Percentage and cp/ml.

| Group | Variable | N Non-Rej. | N Rej. | Sensitivity | Specificity | AUC | PPV at 25% | NPV at 25% |
| --- | --- | --- | --- | --- | --- | --- | --- | --- |
| Validation group | cp/mL | 279 | 75 | 0.71 (0.60-0.78) | 0.91 (0.88-0.94) | 0.89 (0.82-0.95) | 0.72 (0.62-0.81) | 0.90 (0.87-0.93) |
|  | Percent | 279 | 75 | 0.76 (0.67-0.84) | 0.84 (0.81-0.88) | 0.86 (0.77-0.93) | 0.61 (0.54-0.70) | 0.91 (0.88-0.94) |
|  | CM-Score | 279 | 75 | 0.72 (0.61-0.79) | 0.94 (0.92-0.97) | 0.90 (0.83-0.96) | 0.81 (0.72-0.89) | 0.91 (0.88-0.93) |
| Total cohort | Percent | 308 | 106 | 0.76 (0.69-0.83) | 0.85 (0.82-0.89) | 0.88 (0.80-0.93) | 0.63 (0.56-0.71) | 0.92 (0.89-0.94) |
|  | cp/mL | 308 | 106 | 0.72 (0.62-0.77) | 0.91 (0.88-0.94) | 0.89 (0.82-0.94) | 0.72 (0.63-0.80) | 0.91 (0.87-0.93) |
|  | CM-Score | 308 | 106 | 0.72 (0.63-0.78) | 0.95 (0.93-0.97) | 0.91 (0.84-0.95) | 0.82 (0.74-0.90) | 0.91 (0.88-0.93) |

*Rejection vs. Non-Rejection plus Stable; Borderline rejection excluded; NPV and PPV were calculated at 25% prevalence*

### 3.3 Contextualization with Evidence from Other Clinical Cohorts

To fully understand the value of the CM-Score, an additional analysis was performed to compare its established performance characteristics with 11 clinical cohorts from 10 published studies(5, 6, 24-31) that reported percent and/or absolute values, with or without added clinical parameters, in both for-cause and surveillance settings. Larger clinical studies with published sensitivity and specificity using clinical assays were used to calculate predictive values at a rejection prevalence of 25%. A total of 6,536 samples, including 1,690 rejection cases, were included (Supplementary Table S4 and Supplementary Figure S6). Figure 4 shows the results comparing the performance of the selected clinical cohorts(5, 6, 24-31) with the CM-Score.

**Figure 4:**
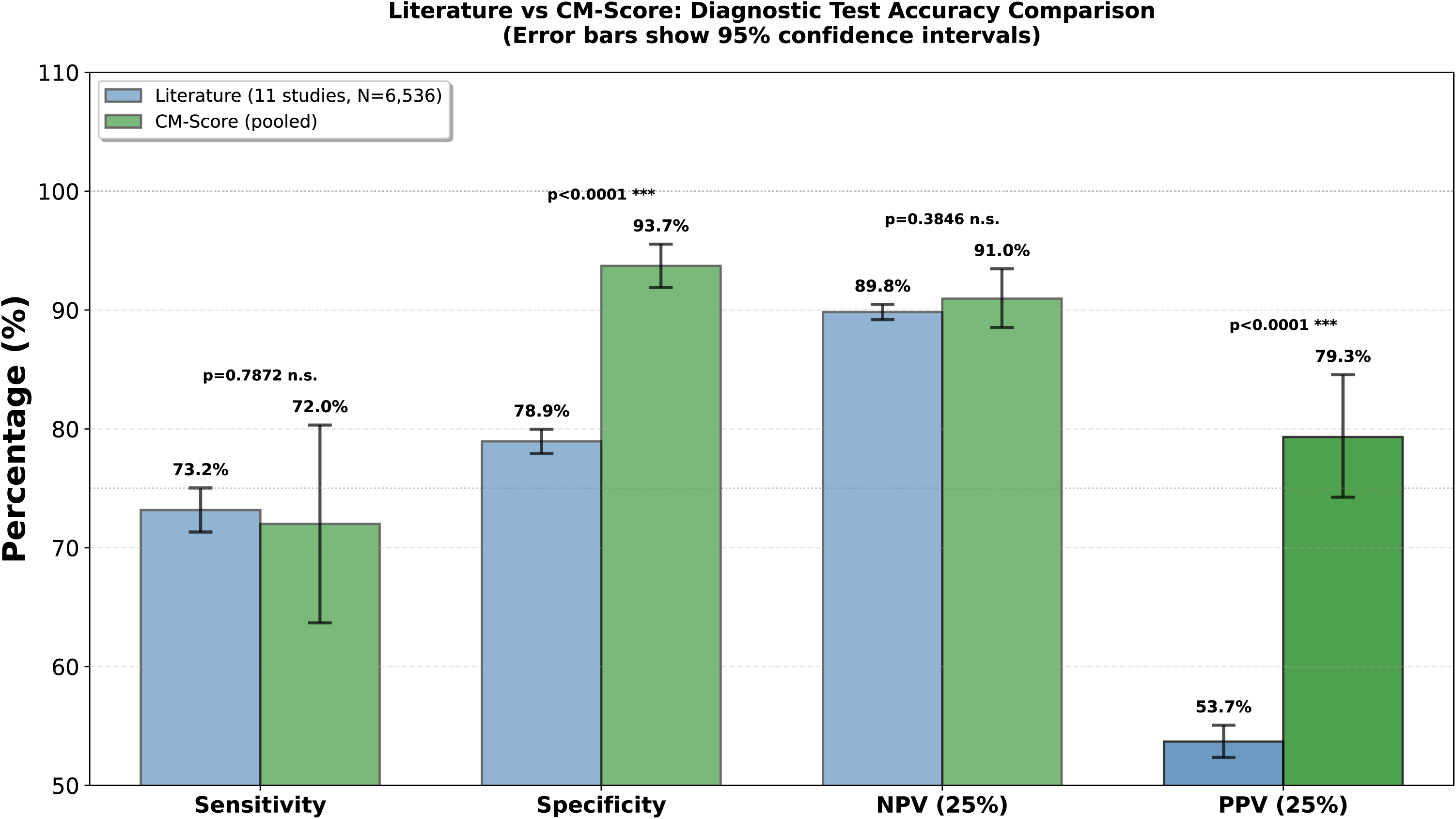
Comparison of the combination of absolute and relative dd-cfDNA (CM-Score) with Data from 11 clinical cohorts from 10 published studies using either absolute or relative dd-cfDNA values. Clinical performance characteristics values and standard deviations are given for published single values compared to the CM-Score in the validation group. Predictive values are calculated at 25% pre-test probability (prevalence) using the published sensitivities and specificities. Details are given in the Supplement.

Combining percent and cp/mL in the CM-Score increased PPV to 81% in the validation cohort while maintaining high NPV, confirming that a continuous integration of both measures outperforms other dd-cfDNA metrics. dd-cfDNA(%) did not differ from the literature data, whereas dd-cfDNA(cp/mL) showed significantly higher specificity, albeit less pronounced than the CM-Score. The full analysis (Supplementary Table S5), shows no differences between the validation group and the full dataset; all subsequent LR analyses used the full dataset. A decision curve analysis comparing the clinical utility of the validation cohort results with published data by quantifying net benefit across a range of threshold probabilities is given in Supplementary Figure S7. Supplementary Table S7 shows the statistical significances of the net benefit of the CM-Score compared to the literature.

### 3.4 Clinical Application

Predictive values are key performance metrics for clinical practice. Calculated via Bayes’ theorem, they depend heavily on prevalence as the pre-test probability. In the for-cause biopsy cohort, the rejection prevalence was 36%, whereas surveillance cohorts typically report rates of 6–11%(32). In general, the ABMR incidence ranges from 3% to 12%(33). In DSA-positive patients, the prevalence of rejection might be up to 40% per year(34, 35), associated with a high PPV(35).

Naesens and Wong recently outlined how dd-cfDNA can be integrated into clinical decision-making(36). Using the likelihood ratios (LR+ and LR–) associated with a test’s defined threshold, the prior probability of rejection can be updated to a post-test probability(37). The general assumption is that an LR^+^ >10 and an LR^-^ <0.33 are considered necessary to confidently rule in or rule out disease. However, these criteria are not met in the published dd-cfDNA cohorts (see Supplementary Table S3).

To support context-specific interpretation, we can use the LRs to exemplify the gain in post-test probability. For the CM-Score (full dataset, N=413) Supplementary Fig. S8 shows the Fagan nomogram with the respective post-test probability, which is a function of the likelihood ratios (LR). Using the single-point cut-off of 0 for the CM-Score and a pre-test probability of 5%, the post-test probabilities are 43% and 1.5%. At 25% pre-test probability, the post-test probabilities are 82% and 8.5%, respectively.

Supplementary Table S8a shows the positive likelihood ratios across the range of expected CM-Score values and pre-test probabilities from 5% to 50%. The corresponding post-test rejection probabilities for CM-Score values exceeding each threshold are also provided. Conversely, Supplementary Table S8b shows the post-test probability when the CM-Score falls below the specified threshold.

LRs can be calculated across the full CM-Score range, providing more reliable, value-based information than a single cutoff. The continuous matrix for distinct CM-Score values is presented in Table 4, providing a more realistic and useful representation of the rejection probabilities. For example, if the CM-Score result is ≥0.8 and the pre-test probability is 35% (suspicious for TCMR, where we can assume equal probabilities), the result would indicate a high post-test probability of rejection of 90%. Conversely, with a very low value of e.g. -0.8, the results would be 1.7%, suggesting that no rejection is present.

**Table 4:** Clinical Interpretation guide for CM-Score.

| Pre↓ \| CM→ | 0.90 | 0.80 | 0.70 | 0.60 | 0.50 | 0.40 | 0.30 | 0.20 | 0.10 | 0.00 | -0.10 | -0.20 | -0.30 | -0.40 | -0.50 | -0.60 | -0.70 | -0.80 | -0.90 |
| --- | --- | --- | --- | --- | --- | --- | --- | --- | --- | --- | --- | --- | --- | --- | --- | --- | --- | --- | --- |
| 5% | 47% | 46% | 45% | 43% | 41% | 38% | 35% | 30% | 25% | 16% | 10.4% | 5.9% | 3.0% | 1.7% | 0.9% | 0.5% | 0.2% | 0.2% | 0.2% |
| 10% | 65% | 64% | 63% | 62% | 60% | 56% | 54% | 47% | 41% | 29% | 20% | 12% | 6.1% | 3.5% | 1.9% | 1.0% | 0.4% | 0.4% | 0.3% |
| 15% | 75% | 74% | 73% | 72% | 70% | 67% | 65% | 59% | 52% | 40% | 28% | 17% | 9.4% | 5.5% | 3.0% | 1.5% | 0.6% | 0.6% | 0.6% |
| 20% | 81% | 80% | 79% | 78% | 77% | 74% | 72% | 67% | 61% | 48% | 35% | 23% | 13% | 7.6% | 4.2% | 2.1% | 0.9% | 0.8% | 0.8% |
| 25% | 85% | 84% | 84% | 83% | 82% | 79% | 78% | 73% | 67% | 55% | 42% | 28% | 16% | 9.8% | 5.6% | 2.8% | 1.2% | 1.1% | 1.0% |
| 30% | 88% | 87% | 87% | 86% | 85% | 83% | 82% | 78% | 73% | 61% | 49% | 34% | 20% | 12% | 7.1% | 3.6% | 1.5% | 1.4% | 1.3% |
| 35% | 90% | 90% | 89% | 89% | 88% | 86% | 85% | 81% | 77% | 67% | 54% | 39% | 24% | 15% | 8.7% | 4.5% | 1.9% | 1.7% | 1.7% |
| 40% | 92% | 91% | 91% | 91% | 90% | 88% | 87% | 84% | 80% | 71% | 59% | 44% | 28% | 18% | 11% | 5.5% | 2.3% | 2.1% | 2.0% |
| 45% | 93% | 93% | 93% | 92% | 92% | 90% | 90% | 87% | 83% | 75% | 64% | 49% | 32% | 21% | 13% | 6.7% | 2.8% | 2.6% | 2.5% |
| 50% | 94% | 94% | 94% | 94% | 93% | 92% | 91% | 89% | 86% | 79% | 69% | 54% | 37% | 25% | 15% | 8.1% | 3.4% | 3.2% | 3.0% |

The same interpretation applies in a surveillance setting, where the probability of rejection is approximately 6-11%. In this context, a CM-Score value of ≥0.8 raises the post-test probability to ≥64%, whereas a negative score of ≤-0.8 reduces it to below 0.5%. Thus, comparison with pre-test probability should guide clinical interpretation: a pronounced increase in the positive post-test probability supports rejection, while a marked decrease argues against it.

Table 4 shows CM-Score thresholds and their dependencies; centers can apply their own experience to estimate local rejection prevalence and derive post-test probabilities to support informed clinical decisions.

Finally, we applied the continuous LR for CM-Score to borderline patients, identifying 11 samples (38%) with a high post-test probability of active rejection (Supplementary Figure 9), which is close to the 33% reported for borderline TCMR as active rejection by molecular pathology(38).

## Discussion

Several publications have demonstrated that adding the absolute quantity of dd-cfDNA to the percentage value improves diagnostic performance in transplant recipients(4, 6, 8). Our results support this finding: at a rejection prevalence of 25%, the CM-Score achieved a PPV of 80.9% in the validation cohort and 82.1% in the full dataset, compared to a weighted average PPV of only 53.7% (52.3%–55.1%) reported in published studies (Figure 4 and Supplementary Table S4). Such low PPVs complicate the interpretation of positive test results. Consequently, dd-cfDNA’s primary clinical utility has been as a rule-out test, supported by consistently high NPVs across studies (weighted average NPV of 89.8%; 89.2%–90.5%) with commercially available assays. However, the low PPV limits utility, as elevated results provide limited guidance for clinical decision-making. The higher PPV achieved by combining percentage and absolute dd-cfDNA into the continuous CM-Score indicates that dd-cfDNA may also have value as a rule-in biomarker. With a PPV of 81-82%, the probability of correctly identifying ongoing rejection is substantially increased, improving confidence in positive results. This improvement reflects the superior performance of the continuous score over simple thresholds or parallel combinations and is supported by a corresponding 168% increase in LR+, indicating markedly stronger rule-in capability. This may be particularly relevant in surveillance settings, where rejection prevalence is lower (6-11%)(32) than in for-cause settings(32). With dd-cfDNA(%) alone or in simple combinations, even when additional clinical predictors are included, positive results often lead to overdiagnosis. Based on published sensitivities and specificities (Supplementary Table S4), the average PPV at 10% prevalence would be only 28%. In current practice, dd-cfDNA replaces surveillance biopsies mainly because of its high NPV, allowing clinicians to forgo biopsies when results fall below thresholds. However, values above the threshold lead to unnecessary biopsies (72%) due to the low PPV. Adding longitudinal data provided limited benefit; Bromberg et al. reported a PPV of 31% at assumed 10% rejection prevalence, within the range of studies without longitudinal data. It is apparent from published clinical cohorts that current dd-cfDNA-based surveillance approaches are associated with a high overdiagnosis ratio, with 2.6 unnecessary biopsies performed for every necessary one (odds of 2.6). Using the dichotomized CM-Score would increase the biopsy yield to 60%, reducing overdiagnosis odds to 0.68 and to ≤0.54 at a high CM-Score of ≥0.8. Furthermore, we show that the CM-Score best discriminates rejection (both TCMR and ABMR) from other pathologies compared to either metric alone. Negative values reliably exclude non-rejection pathologies (e.g., BKVAN, CNI toxicity) with 91% NPV, while positive values predict any rejection type. Although dd-cfDNA, thought to be primarily a damage biomarker, our findings support its relative specificity for rejection. Prior studies demonstrate dd-cfDNA’s relative specificity for rejection (24), particularly in ABMR, with elevated values observed in DSA-negative microvascular inflammation, likely reflecting endothelial injury. The imperfect specificity of around 79% was recently noted by the STAR working group(39). Incorporating both absolute and relative values into the CM-Score improves the specificity for rejection to almost 95%, close to what probably can be achieved with a blood biomarker. It also captures dynamic changes more precisely, potentially enhancing utility for longitudinal and routine monitoring.

Aubert et al. recently demonstrated increased AUCROC in large cohorts when dd-cfDNA was added to a clinical model(24). However, dd-cfDNA percentage alone (AlloSure™/AlloSeq™) showed limited utility, with PPV of 54% at the standard 1.0% threshold and NPV of 86% (Validation group results; calculated at 25% prevalence). When combined with clinical data, NPV increased to 91%, whereas PPV decreased to 52%. Clinical data alone had NPV of 88% and PPV of 50%. This suggests that percentage alone adds marginally to diagnostic value. For surveillance, assuming 10% rejection prevalence, PPV was 22% at the 0.5% threshold and 28% for the 1% threshold. This was confirmed in the Kidney Outcomes with AlloSure Registry (KOAR) study(29), which reported dd-cfDNA >1% or >0.5% with ≥61% relative change (RCV) yielded PPVs of 57% at 25% prevalence (60% using the 1.0% cut-off alone). For surveillance (10% prevalence), PPV drops to 31% and 34%, corresponding to overdiagnosis odds of 2.25 and 1.96, respectively.

It should be understood that focusing solely on AUCROC (a summation value that does not account for curve shape) provides limited information about the added clinical benefit of a diagnostic test(40, 41). The results shown by Aubert et al.(24) exemplify this by demonstrating a significant increase in AUCROC by adding dd-cfDNA without any effect on the PPV. In contrast, we show herein that even without a significant improvement in AUCROC, the clinically important parameters NPV and PPV can still improve substantially and significantly (Fig. 2).

Bromberg et al. reported results from the Prospera Registry (ProActive) study (31), which included mainly surveillance biopsies. At a 25% prevalence, dd-cfDNA(%) yielded an NPV of 93% and a PPV of 64%, corresponding to a PPV of 37% in a surveillance setting. Halloran et al. evaluated the same assay platform in a 218-patient for-cause biopsy cohort, assessing both percentage and copies/mL measurements using a parallel testing rule, in which exceeding either threshold (≥1% or ≥78 cp/mL) constituted a positive result for rejection(6). Compared with Banff-based histology, this approach yielded an NPV of 90% and a PPV of 56% at a 25% prevalence. However, in a surveillance setting, the PPV would decrease to 30%.

Using a two-threshold approach with optimized values (≥0.72% and ≥69 cp/mL) yielded an NPV of 92% (90%–94%) and a PPV of 72% (68% –77%) in our cohort of 106 rejection and 307 non-rejection cases at a 25% rejection prevalence. This improved performance, compared to the same approach reported for NGS-based assays (6), may be due to the use of digital PCR for cfDNA quantification, together with per-sample extraction efficiency measurement. Digital PCR reliability for DNA quantification is well-established and is listed as an example of a primary reference measurement procedure in ISO17511:2020(42). cfDNA extraction from plasma can vary substantially, and controlling for this improves quantification reliability.

Our results confirm that dd-cfDNA percent does not differ from that reported in the literature (Supplementary Table S5b and Figure S6), as expected in a cohort with a wide distribution of time after engraftment(2). In contrast, combining percent and cp/mL into a continuous score performs better than simple thresholds or combinations, as shown by increases in PPV from 72% to 81-82% and in LR+ by 175%.

Furthermore, when incremental discrimination was assessed using bootstrap ΔAUROC, addition of clinical variables (eGFR, time from transplantation, HLA mismatch) to the CM-Score did not result in a meaningful improvement in discrimination, with all ΔAUROC confidence intervals overlapping zero (Supplementary Table 6).

The promising results for resolving borderline TCMR are hypothesis-generating and will be subject to a focused study using molecular pathology as an arbitrator.

The CM-Score offers reliable, standalone diagnostic information; its high PPV and specificity support rejection-directed diagnosis and help distinguish rejection from other causes, such as CNI toxicity, in cases of rising creatinine or falling eGFR.

Table 4 shows the importance of considering the pre-test probability of rejection when interpreting results. The value of using a continuous score rather than dichotomized thresholds, such as the multiple percentage cut-offs proposed by Bromberg et al (29), or the evaluation of percentage and cp/mL described by Halloran et al. (6), is evident. This allows context-dependent interpretation of dd-cfDNA values and is particularly useful in unselected surveillance settings, where rejection prevalence is substantially lower than in for-cause testing. In surveillance, intervention should be limited to clearly elevated results to avoid overdiagnosis; for example, with a 10% baseline rejection rate and a 50% post-test probability biopsy threshold, Table 5 indicates a CM-Score >0.3.

The introduction of the clinical application guide herein, considering the pre-test probability of the event to evaluate the likelihood of the results, allows for a more personalized approach to patient care. It acknowledges that the same result may carry different implications depending on the clinical context and patient’s individual risk profile (e.g. high immunological risk), thereby moving away from a uniform, “one-size-fits-all" interpretation.

In summary, this verification-stage study establishes that the CM-Score has superior diagnostic performance compared to single-metric dd-cfDNA. Prospective clinical validation will be necessary to confirm these findings, verify the impact on clinical decision-making, and evaluate long-term patient outcomes when using the CM-Score to guide biopsy decisions and immunosuppression management.

### limitations

These real-world data may be less controlled but better reflect routine use of dd-cfDNA; although most biopsies were for-cause, Banff-based interpretation is comparable, supporting the use of Bayes’ theorem to estimate post-test probabilities across different pre-test prevalences. Notably, one publication(31) showed slightly improved LRs for surveillance compared to for-cause. Aubert et al. hypothesized that surveillance might result in lower dd-cfDNA values in ABMR(24). Inspection of Figure 2 and Extended Figure 4 reveals roughly twice the dd-cfDNA in all Banff groups in the validation cohort, suggesting methodological bias between the two assays. In our study, central pathology was unavailable, and molecular pathology (MMDx) was limited. Incorporating MMDx could have improved NPV and PPV, given its ∼10% higher concordance(6).

## Data Availability

The data that support the findings of this study are not publicly available, as public data sharing was not included in the ethics approval or patient consent. However, deidentified data underlying the results of this article can be made available upon reasonable request to the corresponding author, provided that such sharing is in accordance with institutional and ethical regulations. Summary statistics and analytic code may also be shared on reasonable request to enable reproducibility of the findings.

## Abbreviations

AUCROC: area under receiver operating curve
ddPCR: droplet-digital PCR
DGF: delayed graft function
DSA: donor-specific antibodies
eGFR: estimated glomerular filtration rate
GN: glomerulonephritis
IFTA: interstitial fibrosis / tubular atrophy
LR: likelihood ratio
MMDx: Molecular Microscope Diagnostic System
MVI: microvascular inflammation
NGS: next-generation sequencing
NPV: negative predictive value
PPV: positive predictive value
RCV: reference change value
ROC: Receiver Operating Characteristic
SNP: single nucleotide polymorphism
SOC: standard-of-care
TCMR: T cell-mediated rejection
UTI: urinary tract infection

## Declaration of Generative AI and AI-assisted technologies in the writing process

During the preparation of this work the authors used ChatGPT (GPT-5, OpenAI) and Claude (Sonnet 4.5, Anthropic) for assistance with language refinement and rephrasing to improve readability. After using this tool, the authors reviewed and edited the content as needed and take full responsibility for the content of the publication.

## Supplementary Material

Supplementary material is available online.

## Author’s contributions

LB, AA, ES, and KB designed the study. LB, AA, JB, AS, KBK, ES, and KB analyzed and interpreted the data and drafted the manuscript. Patients were recruited and results generated by LB, AA, BO, CM, EVS, VS, and KB. JB, ES, THT, MS, and EW established and performed the quantification of dd-cfDNA in their respective laboratories. MO, CM, VS, and KB supervised the project and revised the manuscript. All the authors critically reviewed the manuscript.

