## Supplementary Material for "Improving diagnostic performance of kidney allograft rejection with a model combining relative fraction and absolute copies of donor-derived cell-free DNA - results from five independent cohorts"

**Supplementary Figure S1: Distribution of the time after transplantation**

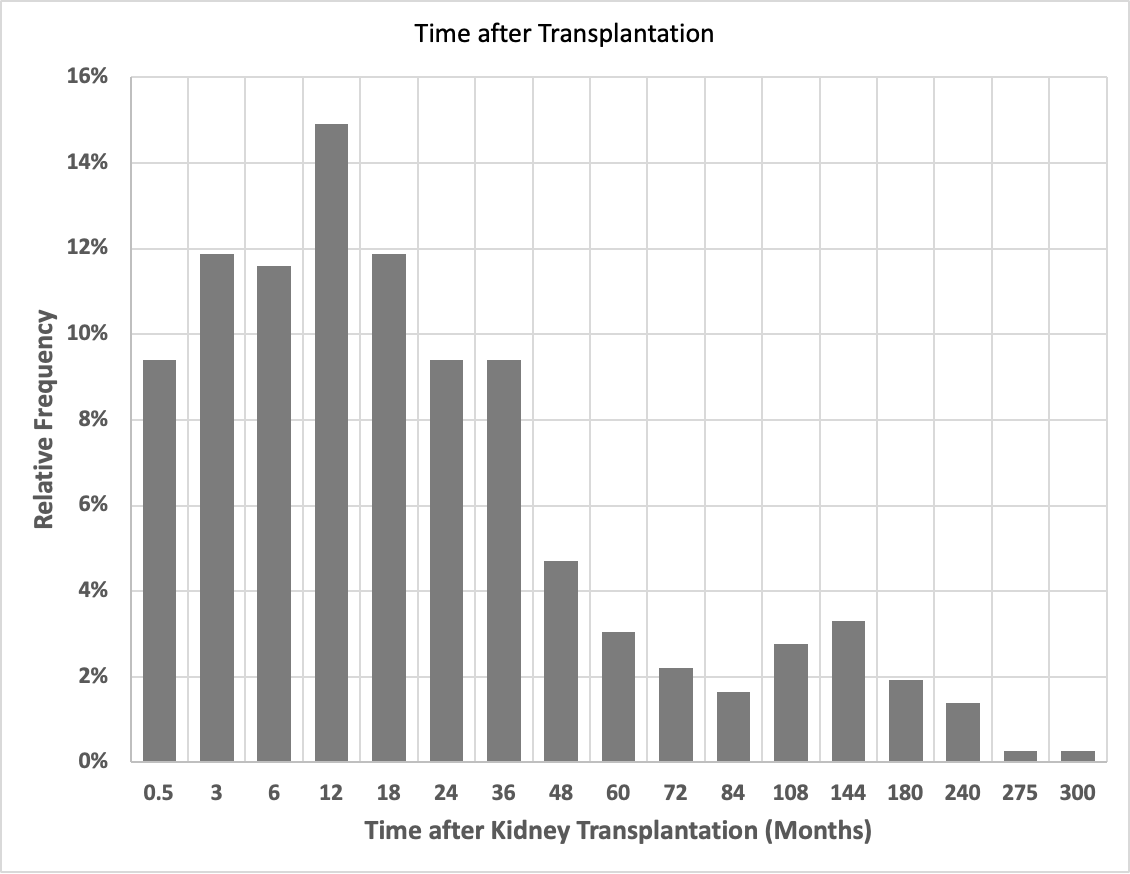

**Supplementary Table S1: dd-PCR methods used in the study**

| Assay Version | Name | Technology | Method |
| --- | --- | --- | --- |
| Generation 1 | VitaGraft Kidney | Bio-Rad QX200 | Genotyping with selected informative assay set |
| Generation 2 | GraftAssure | Bio-Rad QX600 | 45 SNPs interrogated in parallel |

**Figure S2:** **Comparison of Vitagraft (LDT) to the second generation of the assay named GraftAssure.**
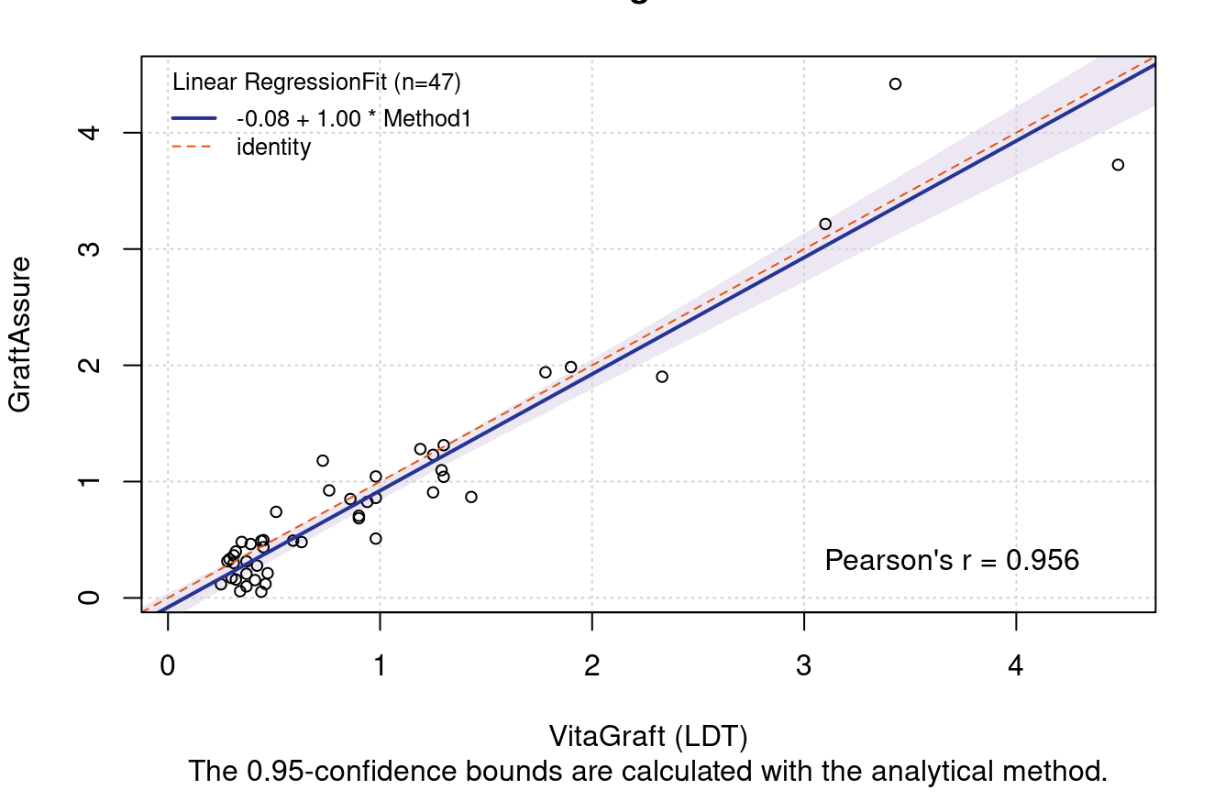

The slope of the regression is 1.00 with confidence limits of 0.91 to 1.09 and an intercept of -0.08% with confidence limit of -0.2% to 0.04%. This shows that both assays will yield values that are statistically identical.

**Supplementary Table S2: Samples used in Discovery and Validation of the Combination Model**

|  | **Discovery** | **Validation** |
| --- | --- | --- |
| **ABMR** | 19 | 41 |
| **Clin. Stable** | 16 | 130 |
| **Neg. Biopsy** | 4 | 21 |
| **TCMR** | 6 | 16 |
| **DSA- MVI** | 5 | 7 |
| **CNI Tox** | 2 | 19 |
| **GN** | 2 | 18 |
| **Mixed Rej** | 1 | 11 |
| **IFTA** | 1 | 21 |
| **other** | 0 | 7 |
| **BKVAN** | 3 | 42 |
| **UTI** | 0 | 6 |
| **ATI** | 1 | 15 |

**Supplementary Figure S3: Correlation between cp/mL and percent (LLoQ to 2 x ULN)**

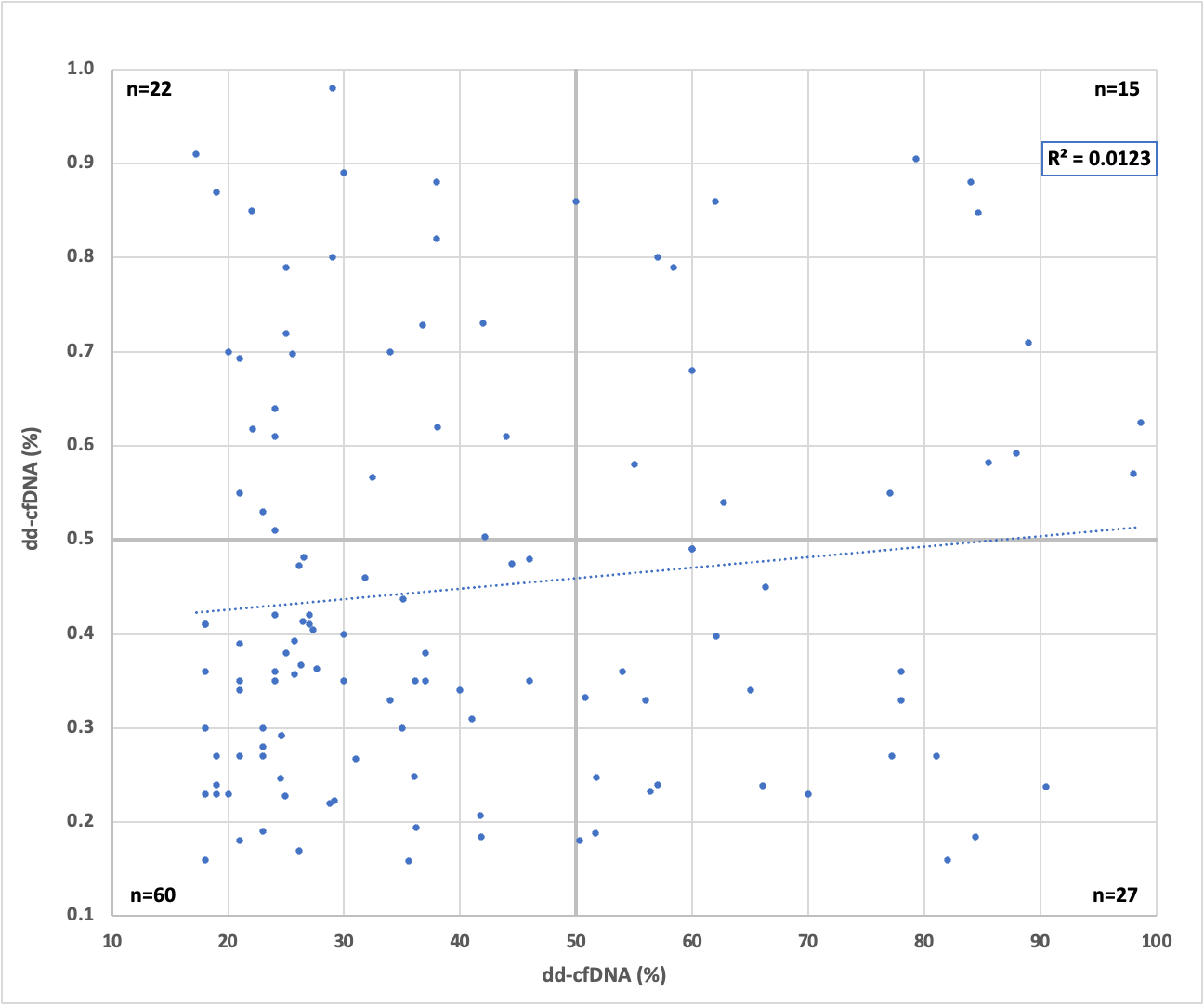

The dispersion of the data shows there is no statistically significant correlation between cp/mL and percentage in the important range (R^2^=0.0123, *P*=0.12). The agreement based on the published thresholds of 0.5% and 50cp/mL^5^ is 60.5% with a Cohen’s κ of 0.092, which indicates that there is virtually no agreement

Bootstrap validation demonstrated minimal optimism (all <0.2%), indicating negligible overfitting. Random splits showed excellent stability (coefficient of variation <3%). External validation and bootstrap methods showed high agreement (all differences <2.6%). These two independent approaches also confirm CM-Score's superior specificity (94.8% vs 84.8%) and PPV (82.5% vs 64.1% at 25% prevalence) over dd-cfDNA percent supporting robust, generalizable performance advantages. The following table and figures show the detailed results, compared to the random Validation group.

**Table S3: Results of Bootstrapping and Random Split vs. Validation group data**

| **CM-Score** | **Validation Group** | **Bootstrap (1000)** | **Random Splits (50)** |
| --- | --- | --- | --- |
| **Sensitivity** | 72.0% (61.0-80.9%) | 71.7% (63.7-80.4%) | 71.7% (71.3-72.1%) |
| **Specificity** | 94.3% (90.9-96.4%) | 94.8% (92.4-97.3%) | 94.8% (94.7-94.9%) |
| **PPV** | 80.8% (71.7-87.3%) | 82.5% (74.6-90.7%) | 82.5% (82.2-82.8%) |
| **NPV** | 91.0% (87.9-93.7%) | 90.7% (87.6-94.1%) | 90.7% (90.6-90.8%) |
| **AUC** | 0.903 (0.883-0.923) | 0.908 (0.876-0.946) | 0.906 (0.904-0.908) |
| **dd-cfDNA (%)** | **Validation Group** | **Bootstrap (1000)** | **Random Splits (50)** |
| **Sensitivity** | 77.3% (66.7-85.3%) | 79.3% (72.1-87.3%) | 78.9% (78.4-79.3%) |
| **Specificity** | 83.9% (79.1-87.7%) | 84.8% (81.1-88.7%) | 84.8% (84.5-85.0%) |
| **PPV** | 61.5% (54.1-68.0%) | 64.1% (56.5-71.8%) | 64.0% (63.7-64.4%) |
| **NPV** | 91.7% (88.2-94.6%) | 92.3% (89.4-95.5%) | 92.1% (92.0-92.3%) |
| **AUC** | 0.861 (0.841-0.881) | 0.876 (0.837-0.922) | 0.872 (0.870-0.874) |
| **dd-cfDNA (cp/mL)** | **Validation Group** | **Bootstrap (1000)** | **Random Splits (50)** |
| **Sensitivity** | 70.7% (59.6-79.8%) | 71.8% (63.2-80.5%) | 71.6% (71.1-72.0%) |
| **Specificity** | 89.2% (85.1-92.4%) | 89.2% (85.9-92.8%) | 89.3% (89.0-89.5%) |
| **PPV** | 68.6% (60.1-75.7%) | 69.6% (60.8-78.6%) | 69.6% (69.1-70.1%) |
| **NPV** | 90.1% (86.7-92.9%) | 90.2% (87.1-93.8%) | 90.1% (90.0-90.3%) |
| **AUC** | 0.892 (0.872-0.912) | 0.886 (0.849-0.924) | 0.886 (0.884-0.889) |

**Figure S4A Differenceplot Bootstrap vs Validation Group results**

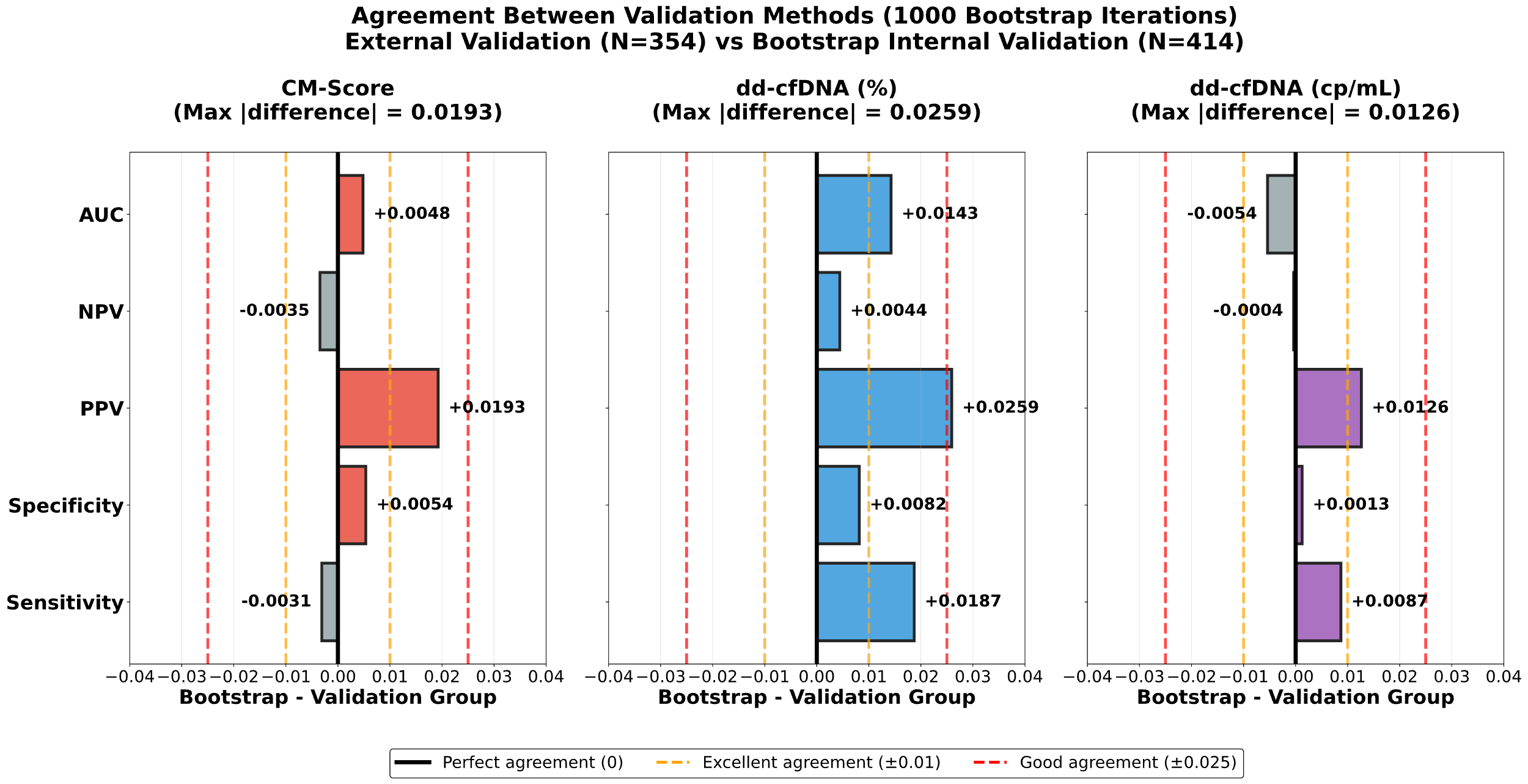

**Figure S4B: Scatterplot of Bootstrap results with Validation Group**

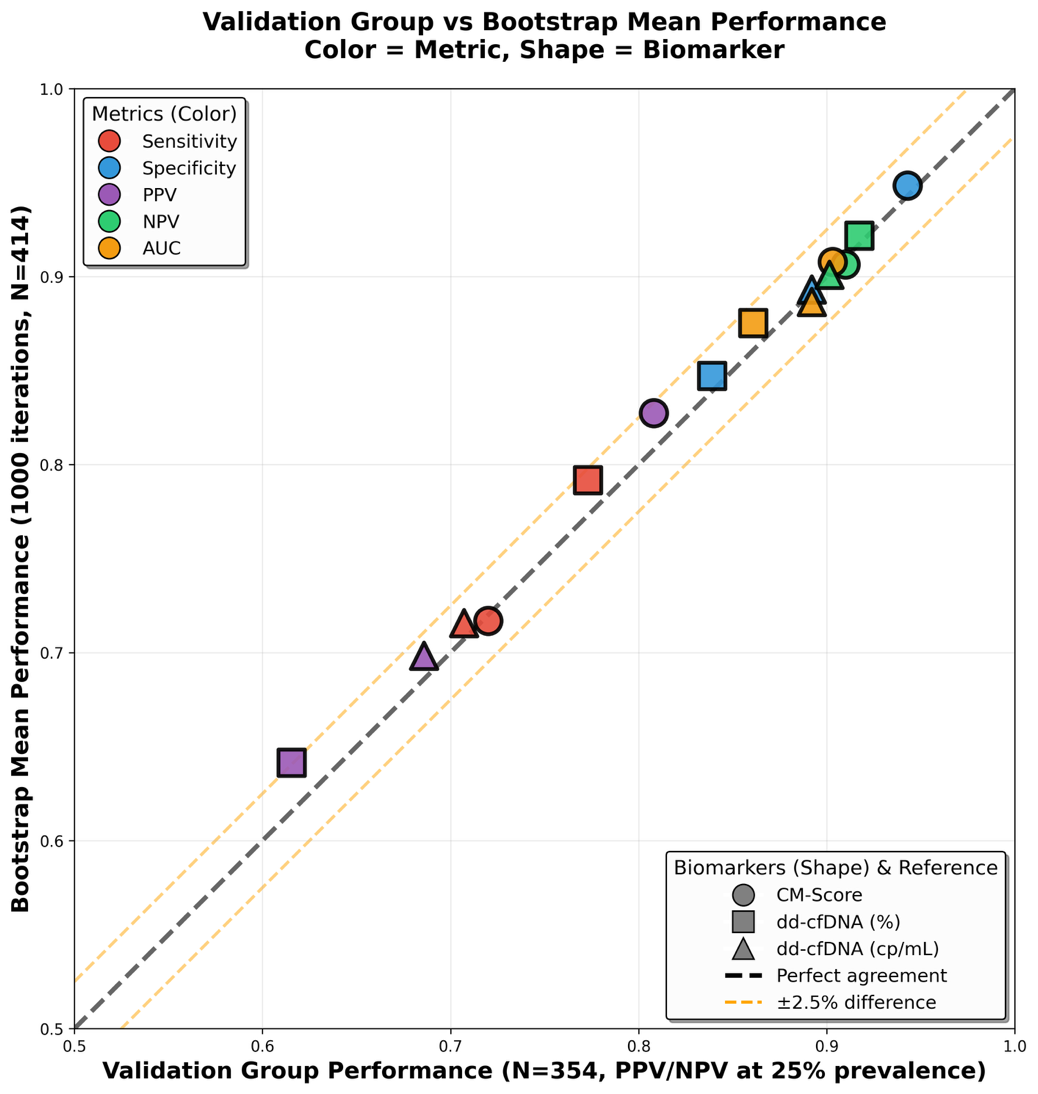

**Supplementary Figure S5 dd-cfDNA in dependence of TCMR severity (TCMR I: n=14; TCMR II: n=8)**

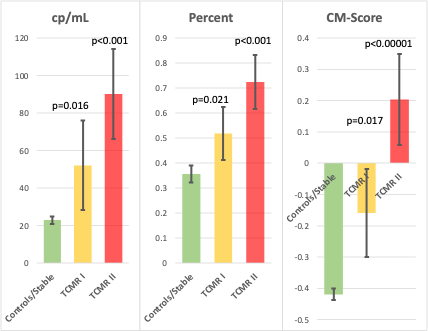

Averages with standard errors are given; *P*-values are vs Control/Stable group.

**Comparison with data from published studies:**

For the calculation of sensitivity and specificity references 6(Halloran), 24(Aubert), 25(Bloom),27(Huang) 28 (Sigdel)and 29(Bromberg1) included borderline TCMR as “Non-Rejections”, whereas the remaining studies excluded borderline TCMR entirely from the calculations

We performed two sensitivity analyses:

1) For the literature 5,6,24-31 there was no statistically significant difference (p>0.05) for sensitivity and specificity between (Table S5a):

All studies vs. studies including borderline(Ref:6,24,25,27-29),

All studies vs. studies excluding borderline (Ref: 5,26,30,31),and

Studies including borderline vs studies excluding borderline.

2) No significant difference was found for specificity when comparing CM-Score with:

borderline included as “Non-Rejection” vs borderline excluded (p>0.424).

Nevertheless, for the comparison of the 11 pooled cohorts from 10 publications, we used CM-score (pooled), for which data were extracted with and without borderline as non-rejection. we calculated borderline proportions separately for rejection and normal samples in the 11 literature studies (1,128/1,690 = 66.8% and 3,541/4,846 = 73.1%respectively), then averaged these to obtain 69.9% borderline and 30.1% no-borderline weighting factors. We applied weighted pooling (weighted value = borderline value × 0.699 + no-borderline value × 0.301) to our CM-Score, cp/mL, and Percent data to match the literature's borderline distribution.

**Supplementary Table S4: Publications used in meta-analysis for comparison of CM-Score with literature**

| **Publication** | **Source** | **Threshold** | **Sensitivity** | **Specificity** | **LR+** | **LR-** | **Samples** | **Rejections** |
| --- | --- | --- | --- | --- | --- | --- | --- | --- |
| **Aubert^24^** | Suppl. Table 6 | Model^1^ | 72.1% | 78.4% | 3.34 | 0.36 | 1,415 | 221 |
|  | Suppl. Table 7 | Model^1^ | 76.2% | 77.7% | 3.42 | 0.31 | 2,317 | 679 |
| **Bromberg^29^** | Figure 3 | Model^2^ | 60.0% | 85.0% | 4.00 | 0.47 | 615 | 118 |
| **Hidalgo^30^** | Table 5(conventional) | 1.0% | 61.8% | 80.1% | 3.11 | 0.48 | 616 | 309 |
| **Bromberg^31^** | Suppl. Table 2 | 1.0% | 79.3% | 85.3% | 5.39 | 0.24 | 249 | 58 |
| **Halloran^6^** | Table 2 (Banff) | 1%\|78cp/mL | 73.5% | 80.8% | 3.83 | 0.33 | 218 | 71 |
| **Bu^26^** | Table 3 | 0.5% | 78.0% | 71.0% | 2.69 | 0.31 | 219 | 106 |
| **Oellerich^5^** | Table 3 | 0.5% | 73.0% | 73.0% | 2.70 | 0.37 | 417 | 22 |
| **Sigdel^28^** | Abstract | 1.0% | 88.7% | 72.6% | 3.24 | 0.16 | 300 | 38 |
| **Bloom^25^** | Page 2223 text | 1.0% | 59.0% | 85.0% | 3.93 | 0.48 | 107 | 27 |
| **Huang^27^** | Figure 4a | 0.74% | 79.4% | 72.4% | 2.88 | 0.28 | 63 | 34 |
|  | Weighted Average | 0.85% | 73.2% | 78.9% | 3.47 | 0.34 | Σ: 6,536 | 1,690 |
|  | CI^95^ Low CI^95^ High | 0.43% 1.27% | 71.3% 75.1% | 77.9%  80.0% | 3.23 3.76 | 0.310.37 |  |  |

1) *Best performing Model incl. dd-cfNDA and “SOC parameters” 2) ≥1% or ≥0.5% and ≥61% increase from previous value.*

**Supplementary Table S5a: Comparison of literature concerning borderline.**

|  | |  | | **All Literature** | | **Literature BL Yes** | |  | | **All Literature** | **Literature BL Yes** |
| --- | --- | --- | --- | --- | --- | --- | --- | --- | --- | --- | --- |
| **Literature BL Yes** | | **Sensitivity** | | Z=+0.94 (p=0.345) | | — | | **Specificity** | | Z=+0.12 (p=0.901) | — |
| **Literature BL No** | |  |  | Z=+0.99 (p=0.325) | | Z=+1.63 (P=0.104) | |  |  | Z=+0.30 (p=0.767) | Z=+036 (P=0.717) |

*Comparison of CM-Score in pooled literature cohort (All) and sub-cohorts with (BL Yes)and without (BL No) borderline TCMR included as non-rejection, revealed no significant differences.*

**Supplementary Table S5b: Performance Metrics (Validation Group) vs All Literature:**

| **Metric** | **Literature (N=6,536)** | **CM-Score  (pooled, weighted)** | **cp/mL  (pooled, weighted)** | **Percent  (pooled, weighted)** |
| --- | --- | --- | --- | --- |
| **Sensitivity, %** | 72.9 (67.8-78.0) | 72.0 (64.4-79.6) Z=-0.20, p=0.8402 | 72.5 (65.0-80.1) Z=-0.09, p=0.9313 | 77.9 (70.9-84.9) Z=1.11, p=0.2655 |
| **Specificity, %** | 78.9 (76.2-81.6) | 93.3 (91.1-95.5) **Z=8.13, p<10^-15^** | 86.9 (83.9-89.8) **Z=3.90, p<0.0001** | 80.2 (76.7-83.6) Z=0.58, p=0.5610 |
| **PPV at 25%, %** | 53.5 (49.9-57.1) | 78.2 (72.3-84.0) **Z=7.00, p<10^-11^** | 64.8 (59.1-70.4) **Z=3.29, p=0.0010** | 56.7 (51.9-61.5) Z=1.04, p=0.2996 |
| **NPV at 25%, %** | 89.7 (88.0-91.5) | 90.9 (88.7-93.2) Z=0.80, p=0.4234 | 90.5 (88.1-92.8) Z=0.48, p=0.6299 | 91.6 (89.1-94.0) Z=1.19, p=0.2356 |
| **LR+** | 3.45 (2.95-3.96) | 10.73 (7.04-14.42) **Z=3.83, p=0.0001** | 5.52 (4.15-6.88) **Z=2.78, p=0.0055** | 3.93 (3.16-4.70) Z=1.01, p=0.3113 |
| **LR-** | 0.34 (0.28-0.41) | 0.30 (0.22-0.38) Z=-0.80, p=0.4221 | 0.32 (0.23-0.40) Z=-0.48, p=0.6291 | 0.28 (0.19-0.36) Z=-1.19, p=0.2327 |

**Supplementary Table S5c: CM-score performance metrics in different groupings**

| **Group** | **N** | **Sensitivity** | **Specificity** | **PPV** | **NPV** | **LR+** | **LR-** |
| --- | --- | --- | --- | --- | --- | --- | --- |
| CM-Score Validation Group (without BL) | 354 | 72.0% | 93.9% | 79.7% | 91.0% | 11.80 | 0.3 |
|  |  | (67.3%–76.7%) | (89.2%–98.6%) | (67.2%–92.2%) | (89.5%–92.4%) | (2.68–20.93) | (0.25–0.35) |
| CM-Score All (pooled*) with/without BL | 428 | 73.6% | 93.9% | 80.1% | 91.4% | 12.07 | 0.28 |
|  |  | (68.2%–79.0%) | (92.2%–95.5%) | (75.6%–84.5%) | (89.8%–93.1%) | (7.74–19.02) | (0.20–0.36) |
| CM-Score All  without BL | 415 | 73.6% | 94.5% | 81.6% | 91.2% | 13.34 | 0.28 |
|  |  | (68.2%–79.0%) | (91.3%–96.5%) | (74.5%–88.3%) | (88.5%–93.6%) | (8.44–21.89) | (0.20–0.37) |

*95% confidence intervals in brackets *) used for all comparisons with pooled literature data.*

**Visualization of Literature comparisons**

A bubble plot was generated to provide a by study overview of the cited study populations, which visualizes the performance metrics of each study separately together with the pooled data for percent, cp/mL and CM-Score in the herein used population of kidney recipients. It is obvious that the percentage values alone are not different from the studies. (The slight significance seen (Table 5b) is an effect of the narrow Cis of the pooled literature due to the large number of subjects and shall not be overinterpreted). The main improvement is seen with the CM-Score, yielding a widely outstanding PPV due to the improved sensitivity of the combination of percent and cp/mL dd-cfDNA. The effect on both, PPV and NPV over a broad range of prevalences is also shown.

**Supplementary** **Figure S6a: Bubble Diagrams for Sensitivity vs Specificity and NPV vs PPV**

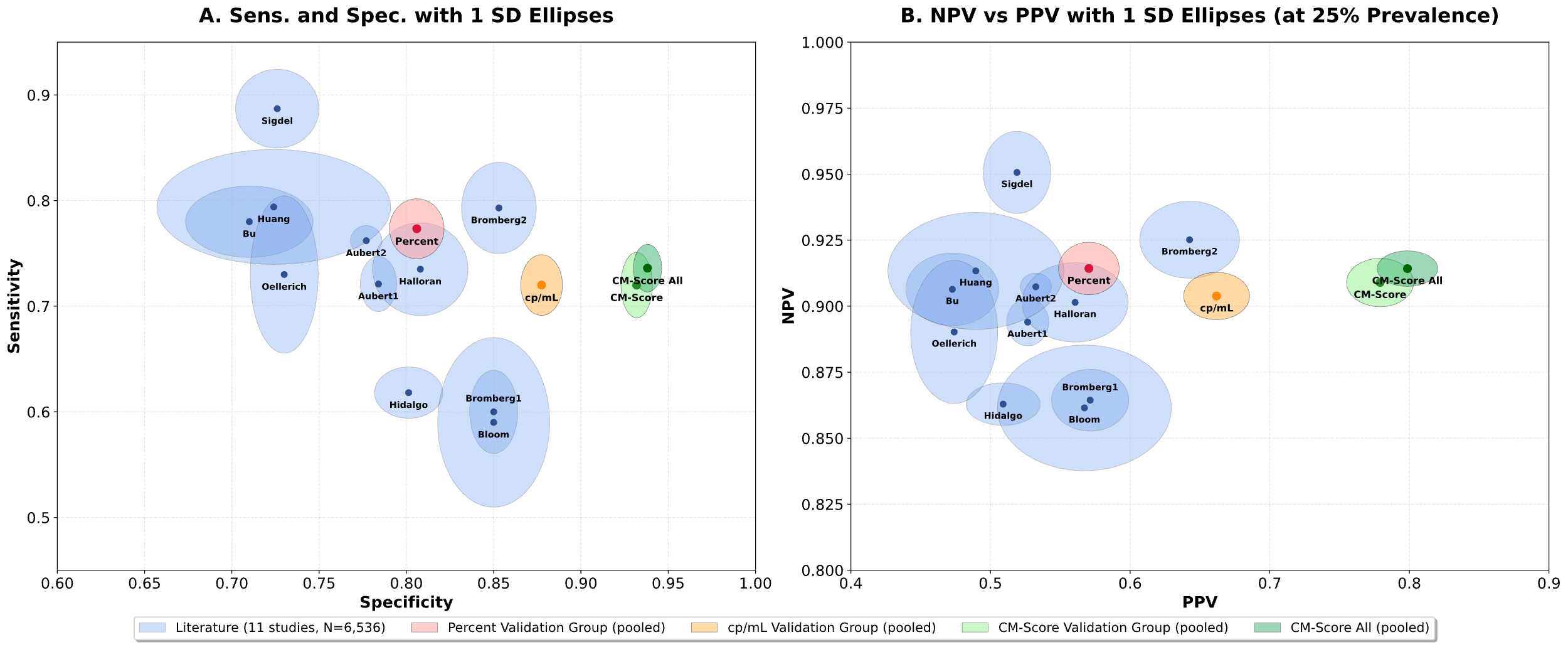

*Aubert1: Discovery cohort; Aubert2: External cohort; Bromberg1: Reference ^29^; Bromberg2: Reference ^31^. For reasons of comparability all GraftAssure data (GA) including CM-score are pooled from data with and without borderline rejections (see above).*

**Supplementary** **Figure S6b: Comparison of PPV and NPV by Prevalence**

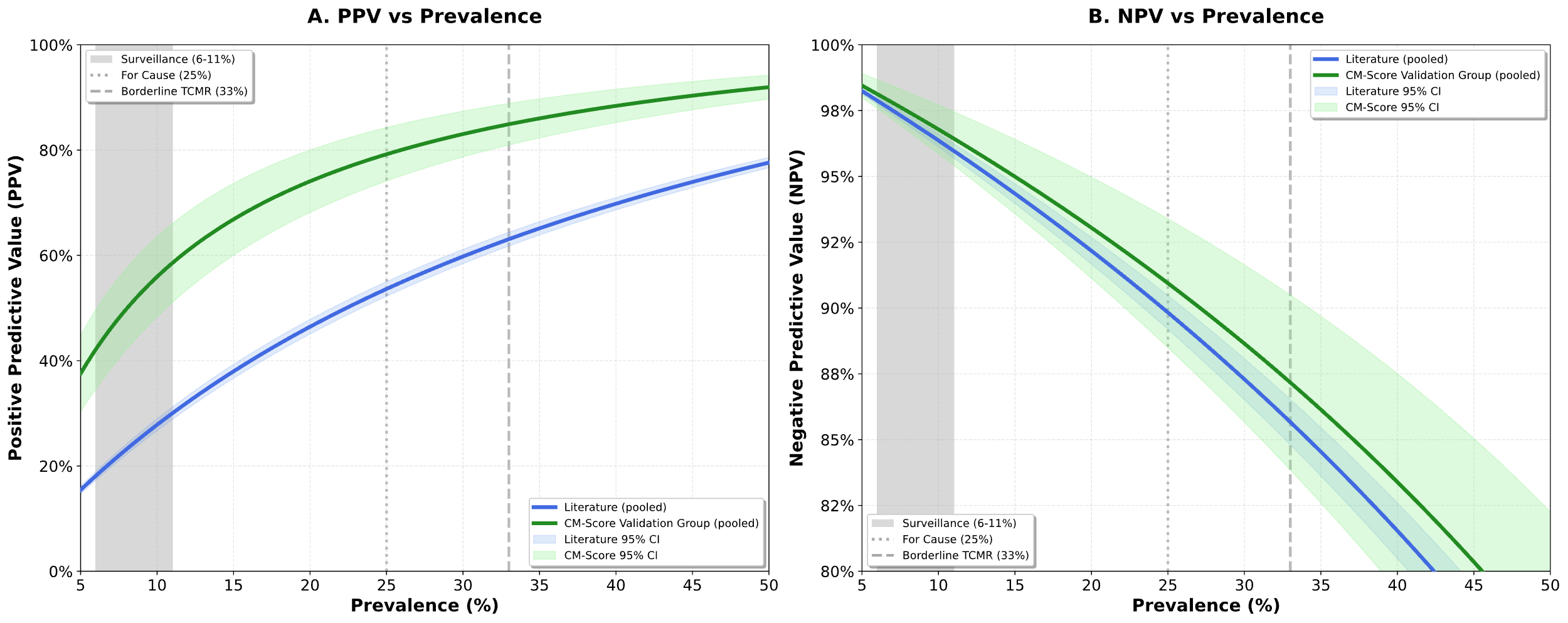

*PPV (A) and NPV (B) as functions of prevalence (5%-50%) for pooled literature (blue, 11 studies, N=6,536) and CM-Score Validation Group (green, N=737). Prevalence references point are given for different clinical scenarios. CM-Score shows superior PPV across all prevalence value. Data from Table S5b were used.*

**Decision Curve Analysis**

Decision curve analysis (DCA) evaluates the clinical utility of a diagnostic test by quantifying the net benefit of using the test to guide treatment decisions across a range of threshold probabilities. The threshold probability represents the minimum probability of disease at which a clinician would choose to intervene (e.g., perform a biopsy, where an aggressive strategy might use a 10% threshold and a conservative strategy would only do a biopsy at greater than 50% threshold). The Net benefit is calculated as the true positive rate minus the false positive rate weighted by the odds of the threshold probability: [TP rate - FP rate × pt/(1-pt)], where pt is the threshold probability. This metric balances the benefit of correctly identifying disease against the harm of unnecessary interventions. The decision curve plots net benefit (y-axis) against threshold probability (x-axis), comparing diagnostic strategies against two reference approaches: "treat all" (intervene regardless of test results) and "treat none" (never intervene). A clinically useful test should demonstrate higher net benefit than both reference strategies across clinically relevant threshold probabilities. Unlike traditional accuracy metrics, DCA directly addresses whether using a test improves clinical decision-making by incorporating disease prevalence and the clinical consequences of false-positive and false-negative results into a single, interpretable framework that reflects real-world decision contexts.

**Supplementary Figure S7: Decision Curve Analysis at different assumed prevalences**

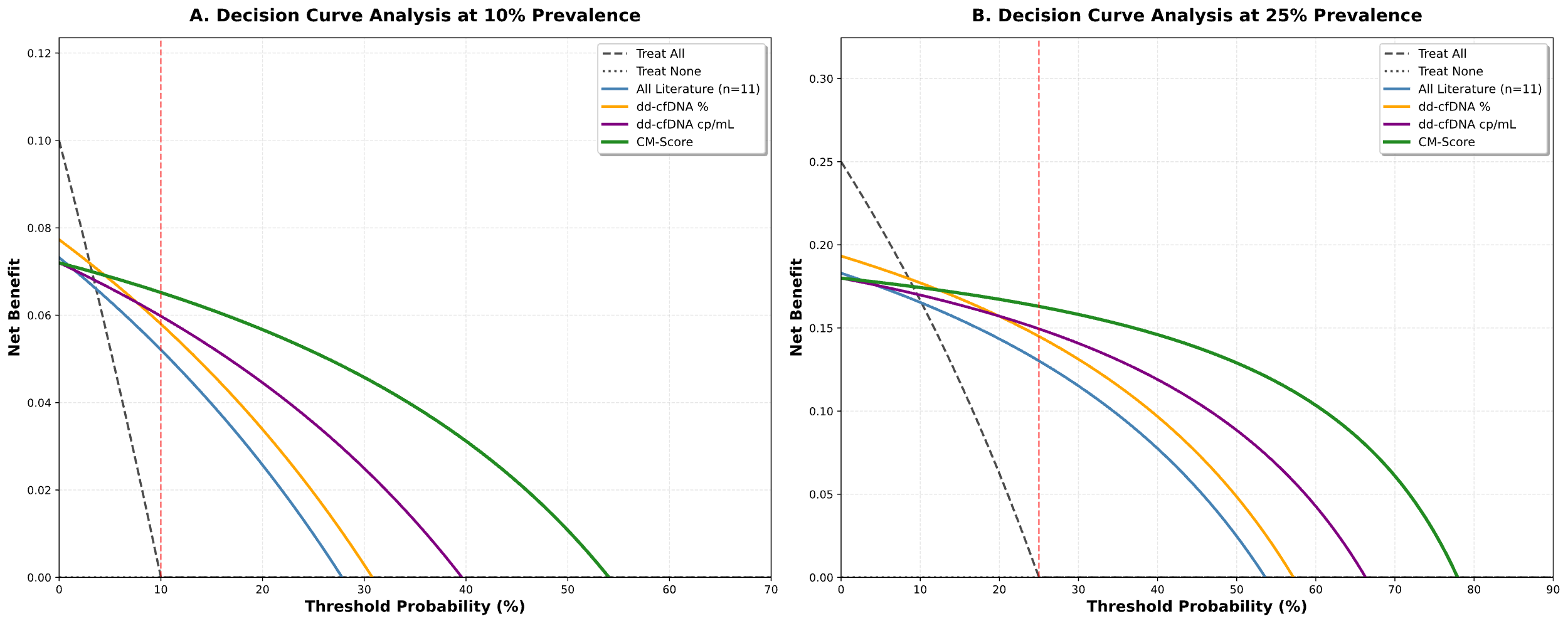

*Net benefit of dd-cfDNA as Percent, cp/mL and CM-Score (including a curve for all values in the study) compared to data from the literature (Suppl. Table S4). The interpretation would be for indication of doing a biopsy.*

**Supplementary Table S6: Pairwise comparison of Thresholds of Decision Curve**

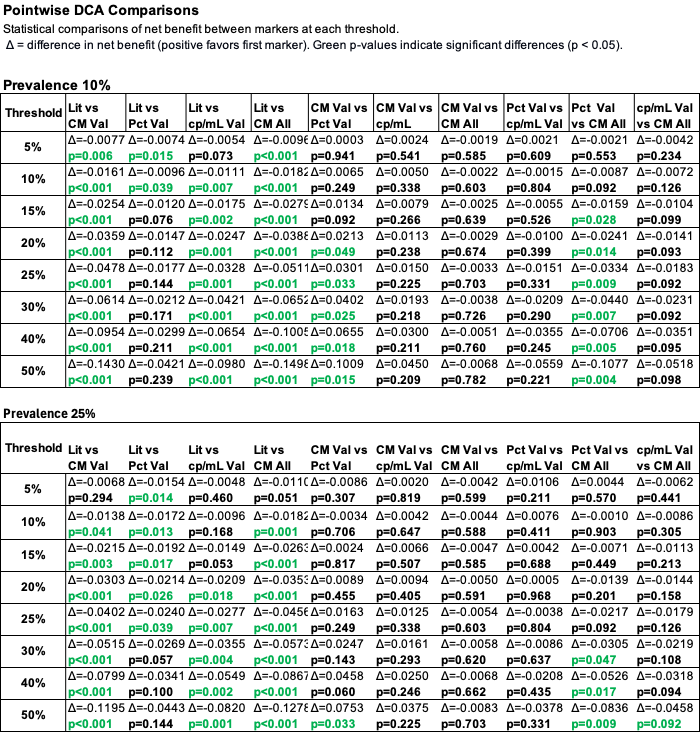

**Validation Group vs. full cohort**

A comparison of the Validation cohort (N=354) with the complete cohort (N=413) revealed no significant differences. Sensitivity: P=0.722; specificity: P=0.823. For the subsequent calculations of continuous LR we therefore used the entire cohort to ensure most robust results especially at the extreme values. The LR+ at the cut-off of SC-score = 0 was 13.34 the LR- was 0.28 (see Table S5c), which yields a PPV of 81.8% (75.7-88.4) and a NPV of 91.5% (89.3-93.7) when the entire cohort was used.

**Addition of clinical data to the CM-Score**

We further investigated if the addition of certain clinical data commonly used in clinical kidney transplant models to predict rejection for their additional value in addition to the CM-score. Data were available for 101 rejections and 230 biopsy proven non-rejections. eGFR, proteinuria, HLA-mismatch and time after transplantation were evaluated for their added value to the AUCROC calculated for the CM-Score alone. No collinearity with the CM-Score was seen (R^2^ <0.065). The difference to the CM-Score AUCROC was marginal and not significant (Table S6a), since the confidence intervals, calculated by bootstrapping did not exclude zero (0). Logistic regressions of the clinical variables with rejection status revealed a slightly significant association with eGFR and a highly significant association with CM-Score (Table 6b). This can explain the dominance of the CM-Score in the multivariate evaluation and the suppression of all other varables added in the AUCROC analyses.

**Supplementary Table S7a: Additional value of clinical Parameters**

| **Model** | **Predictors Added to  CM-Score** | **ΔAUCROC** | **95% Bootstrap CI** |
| --- | --- | --- | --- |
| Reference | CM-Score only | 0 | Reference |
| Model 2 | + eGFR | 0.013 | −0.018 to +0.046 |
| Model 3 | + eGFR + Proteinuria | 0.007 | −0.024 to +0.041 |
| Model 4 | + eGFR + Proteinuria + Time | 0.011 | −0.020 to +0.044 |
| Model 5 | + eGFR + Proteinuria + Time + HLA MM | 0.012 | −0.022 to +0.047 |

*Data from a subset of 331 patient samples, where the clinical parameters were available.2,000 Bootstraps were used. No significant improvement could be detected by adding to the dd-cfDNA CM-Score.*

**Supplementary Table S7b: Logistic correlation with rejection status**

| **Time after Tx** | **HLA-MM** | **Proteinuria** | **eGFR** | **CM-Score** |
| --- | --- | --- | --- | --- |
| F=0.86  P=0.3546 | F=0.23 P=0.6316 | F=0.88 P=0.3488 | F=4.183 P=0.0416 | F=16.683 P=0.0001 |

*F=values with the respective P=values are given. Tx=transplantation, MM=mismatch*

**Supplementary Figure S8: Fagan Nomogram for two different scenarios 5% and 25% pre-test probability (prevalence) using the LRs (positive LR=13.4; negative LR=0.28) from the single cut-point of 0 for the CM-Score.**

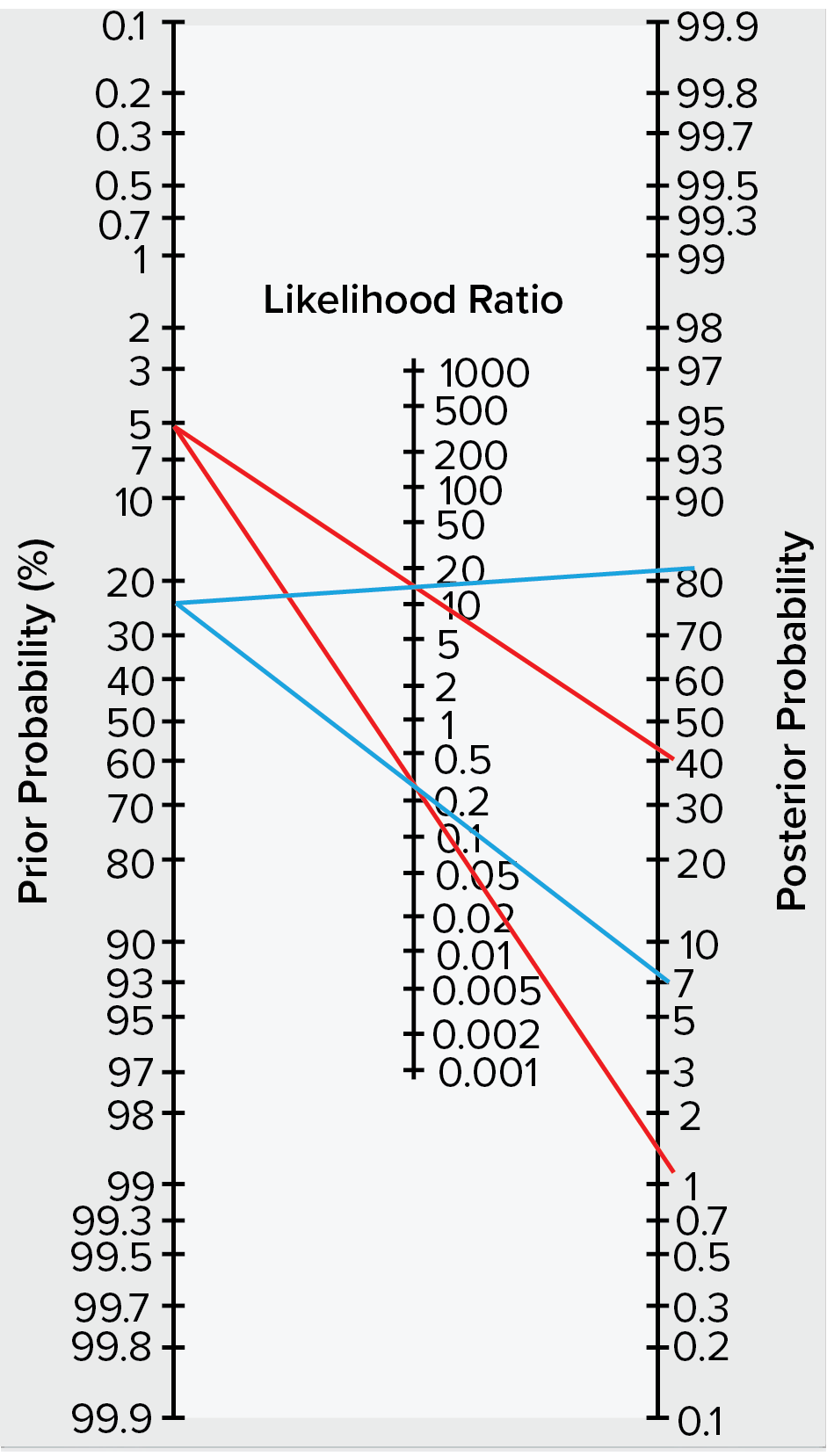

Prior probability: **5%** Posterior probability+= **41%** Posterior probability -= **1.5%**
Prior: probability **25%** Posterior probability += **82%** Posterior probability -= **8.5%.**
The positive probability is for results above the threshold of 0 and the negative probability is for results below this threshold

**Supplementary Table S8: Positive and negative Likelihood ratios of the CM-Score with the calculated post-test probability of rejection stratified by different pre-test probabilities.**

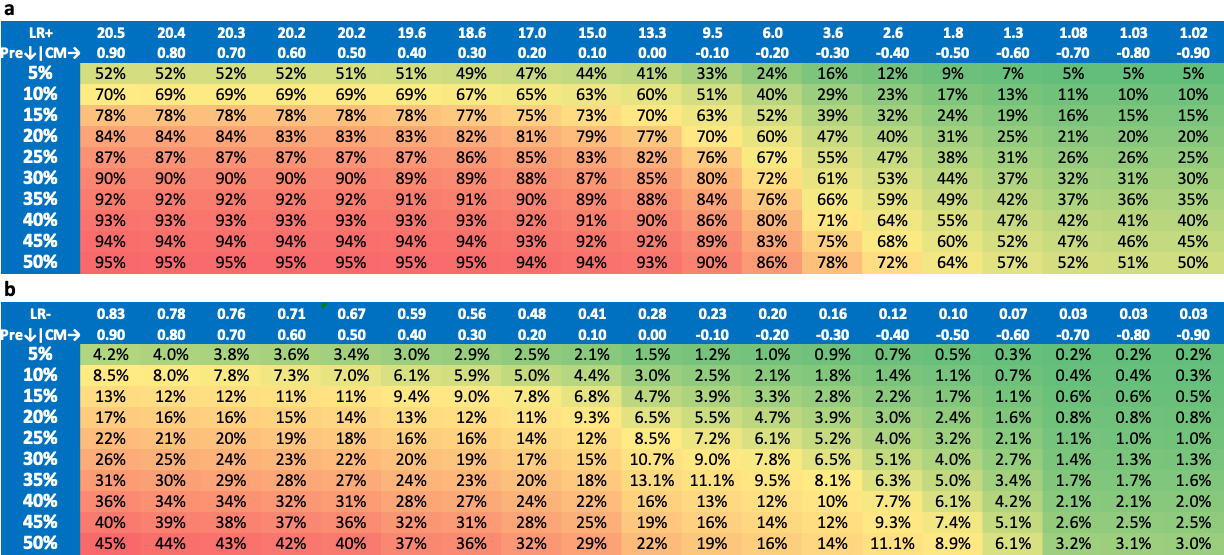

a) Post-test probability of rejection based on the positive likelihood ratio
b) Post-test probability of rejection based on the negative likelihood ratio.

**Borderline TCMR**

Suspicious TCMR samples cannot be discerned into rejection or non-rejection by light microscopy, they are more often called borderline. For this situation of an ambiguous biopsy result dd-cfDNA can aid to apprehend whether a borderline TCMR needs to be considered to benefit from treatment. Here, we assume the pre-test probability with about 33%, based on earlier published molecular pathology findings (Ref. 37). Figure S9 shows that 11 patient samples have a highly elevated post-test probability above the error range of the of the calculations. This results in a fraction of about 38% (19%-57%), compared to the 33% reported(Ref. 37).

**Supplementary Figure S9: Post-test probability for rejection of the group of borderline TCMR**

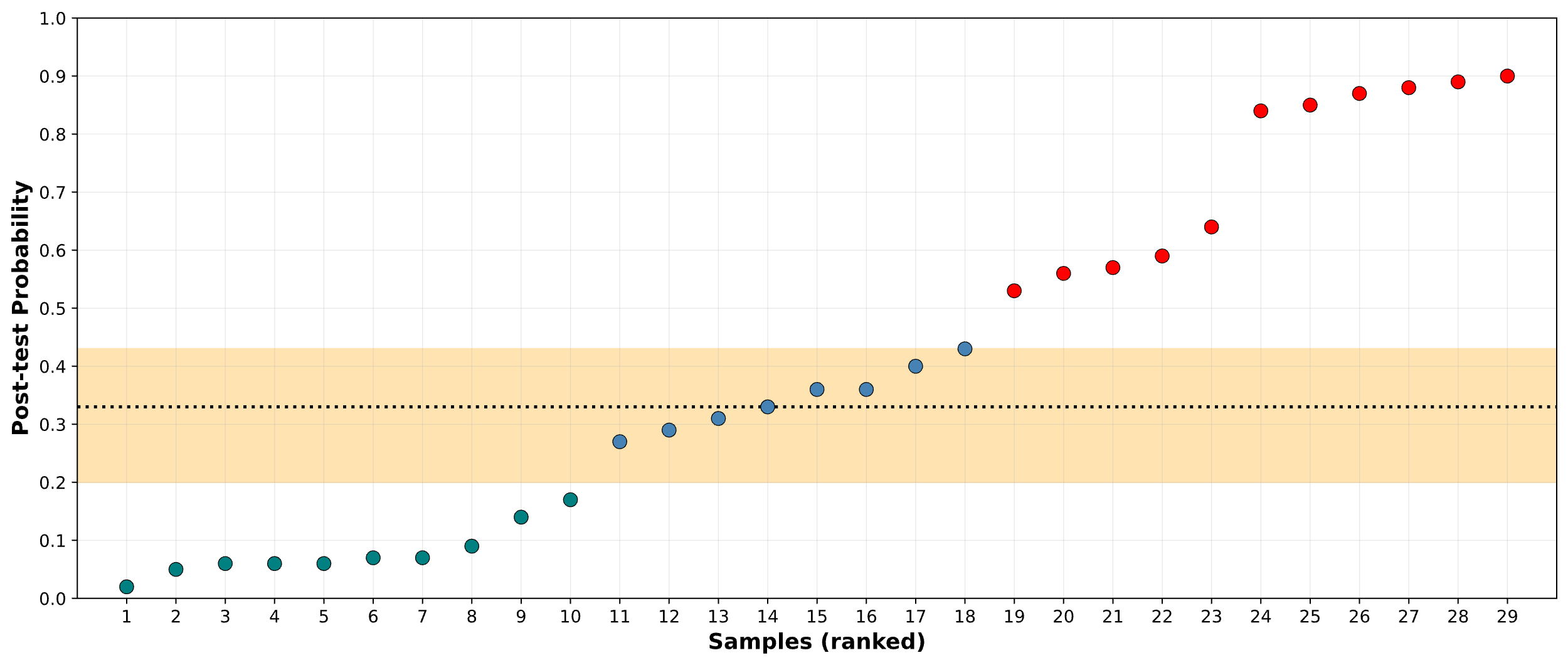

*The dotted line represents the pre-test probability of 33%. The shaded area represents the 95% CI at a LR of 1.0; samples in this area are considered ambiguous for rejection/no rejection. Samples above (red dots) show a high, samples below (green dots) show a low post-test probability.*
